# Non-invasive aspergillosis following COVID-19 exacerbates the severity of SARS-CoV-2 infection

**DOI:** 10.1101/2025.07.03.25330861

**Authors:** Jong Seung Kim, Jun Hyung Park, Juhyun Kim, Ji Won Yang, Wankyu Kim, Yong Chul Lee, Jae Seok Jeong

**Affiliations:** Research Center for Pulmonary Disorders, Research Institute of Clinical Medicine of Jeonbuk National University-Biomedical Research Institute of Jeonbuk National University Hospital, Jeonju, Republic of Korea; Department of Otorhinolaryngology-Head and Neck Surgery, Jeonbuk National University Medical School, Jeonju, Republic of Korea; Department of Medical Informatics, Jeonbuk National University Medical School, Jeonju, Republic of Korea; Respiratory Drug Development Research Institute, Jeonbuk National University Medical School, Jeonju, Republic of Korea; Department of Life Sciences, Ewha Womans University, Seoul, Republic of Korea; Laboratory of Respiratory Immunology and Infectious Diseases, Korea Zoonosis Research Institute, Jeonbuk National University, Iksan, Republic of Korea

**Keywords:** COVID-19, Respiratory aspergillosis, Transcriptome

## Abstract

**Background:** A limited number of large-scale population-based studies regarding the causality between COVID-19 and respiratory aspergillosis exist. Herein, using nationwide data, we investigated whether SARS-CoV-2 infection increases incidence of respiratory aspergillosis and impact of COVID-19-associated aspergillosis on severity of COVID-19. Further, to assess the biological impact of SARS-CoV-2 infection on airway structural and immune cells, we analyzed publicly available COVID-19 transcriptomic datasets.

**Study Design and Methods:** Utilizing a nationwide cohort of 8.5 million clinical registries, we included over 550,000 patients diagnosed with COVID-19 between October 8, 2020, and December 31, 2021, along with control-matched group. The primary outcomes were aspergillosis incidence, including both invasive and non-invasive forms, and its impact on the severity of COVID-19.

**Results:** COVID-19 was closely associated with increased incidence of subsequent respiratory aspergillosis. Comorbidities, including diabetes and COPD, increased the incidence of fungal infections in COVID-19 patients. Regarding severity of COVID-19, both invasive and non-invasive aspergillosis exacerbated the severity of the disease. Particularly, systemic corticosteroids had an overwhelming impact on the increased severity and mortality in both forms of aspergillosis. Notably, antifungal-related genes and pathways, including CCR6, CXCL9, and CX3CR1, were consistently downregulated following SARS-CoV-2 infection and/or corticosteroid treatment.

**Interpretation:** Our findings indicate that COVID-19 increases the incidence of respiratory aspergillosis. Moreover, respiratory aspergillosis, irrespective of its clinical invasiveness, significantly exacerbates the severity of COVID-19. Well-designed studies on the therapeutic potential of antifungal agents to improve the outcomes of COVID-19 are warranted.

## Introduction

Fungi are ubiquitous in daily life and often infect patients with various immune states. *Aspergillus fumigatus (Af)*, a saprotrophic fungus dominant in indoor and outdoor environments, imposes a considerable impact on the respiratory system.^1^ In modern medicine, the widespread use of immunomodulatory agents has increased the incidence and mortality of invasive aspergillosis, especially in immunocompromised patients.^2^ Therefore, invasive aspergillosis poses a substantial threat to public health, as exemplified by the marked rise in mortality.^3,4^ Moreover, even in immunocompetent hosts, accumulating evidence has demonstrated an increasing incidence of aspergillosis, posing a significant burden on public health.^5,6^

In the era of coronavirus disease 2019 (COVID-19), mounting studies have shown an increasing incidence of mycosis associated with viral infections.^7^ In particular, invasive aspergillosis, such as COVID-19-associated pulmonary aspergillosis (CAPA), has been increasingly reported in patients with severe COVID-19.^4^ Although the epidemiological impact of viral infections on the incidence and severity of fungal diseases was repeatedly reported during the 2009 influenza A virus pandemic, the causality between COVID-19 and invasive aspergillosis has yet to be clearly elucidated on a nationwide basis.^5^ Furthermore, whereas studies evaluating CAPA are accumulating, the outcomes of COVID-19-associated respiratory non-invasive aspergillosis (CARNIA) remain unclear, and there is a paucity of large-scale, nationwide population-based cohort research on the impact of non-invasive aspergillosis on patients infected with severe acute respiratory syndrome coronavirus 2 (SARS-CoV-2). Recently, we performed preclinical mechanistic studies using animal models of non-invasive aspergillosis and demonstrated the impact of this condition on the increased severity and mortality in COVID-19.^8^

Using nationwide data derived from the largest population cohort available to date, we focused on whether SARS-CoV-2 infection increases the incidence of respiratory aspergillosis. Moreover, we aimed to provide insights into the impact of COVID-19-associated aspergillosis on the severity of COVID-19 with a particular focus on CARNIA. Importantly, to further validate our epidemiological findings, we investigated whether SARS-CoV-2 infection and associated viral-targeted corticosteroid therapy could influence the antifungal defense of the host using a publicly available COVID-19 dataset.

## Materials and Methods

### Study Design and Data Source

In the primary analysis, the cohort was divided into the COVID-19 and matched control groups to assess the presence (primary outcome) and severity (the other primary outcome) of aspergillosis. This study used data from the National Health Insurance Service-National Health Information Database (NHIS-NHID) (NHIS-2022-1-623). Nearly every person in the Republic of Korea is enrolled in the National Health Insurance System, thus providing access to comprehensive nationwide COVID-19 medical data. The NHIS database includes demographic data, such as age, sex, economic status, and residential area, as well as clinical data including medical records, diagnoses, prescriptions, and medication information. The NHIS-NHID database encompasses detailed records of approximately 580,000 confirmed SARS-CoV-2 cases and about 7.9 million individuals in the control group, covering the period from 2020 to 2021. This dataset integrates demographic information, comorbidity profiles, and medical histories collected between 2015 and 2019. All data were anonymized to ensure privacy protection.

### Study Population and Operational Definitions

The study period was from October 8, 2020, to December 31, 2021 (Figure 1). The COVID-19 group included individuals with laboratory-confirmed COVID-19 and those diagnosed with the International Classification of Diseases, 10th Revision (ICD-10) code U07.1 during this period. The control group comprised individuals matched in a 1:1 ratio with the COVID-19 group, using propensity score (PS) matching to ensure similar demographics and medical histories. The primary outcome was the occurrence of aspergillosis, and the other primary outcome included the following severity indicators: 1) admission to the intensive care unit (ICU), 2) extracorporeal membrane oxygenation treatment, 3) intubation or mechanical ventilation, 4) death, and 5) oxygen supply. Follow-up was conducted until the occurrence of the aforementioned outcomes, death, or the study end date (December 31, 2021), whichever came first.^9^ Systemic steroid therapy within 2 weeks after SARS-CoV-2 infection included intravenous dexamethasone or oral prednisolone administration.

**Figure. 1.**
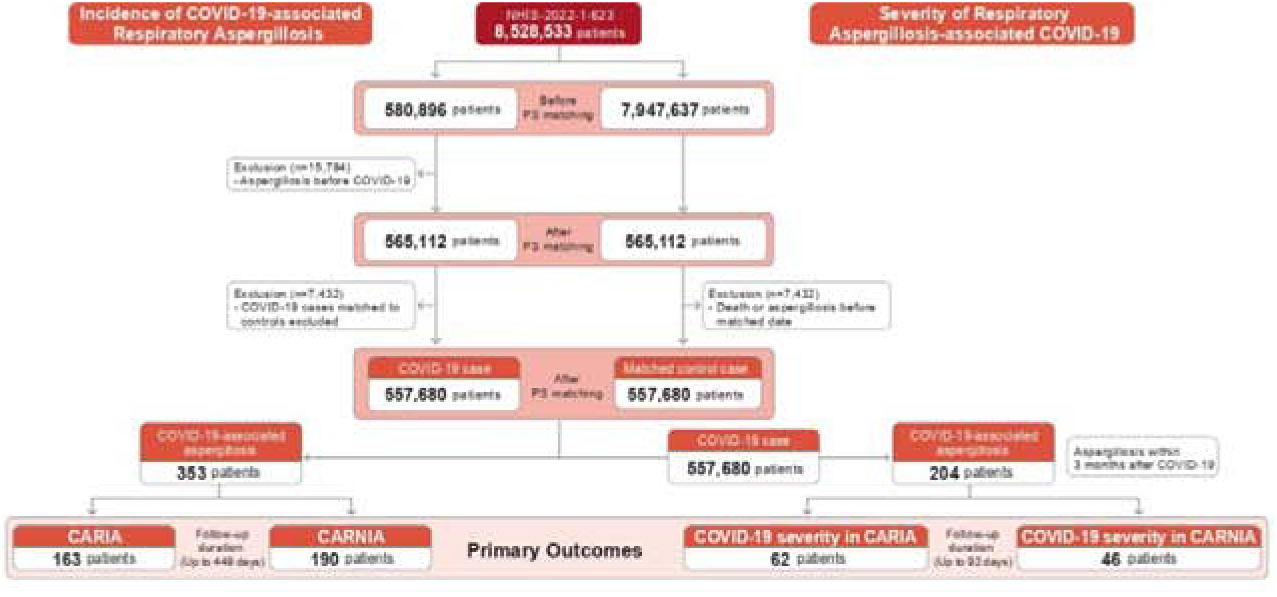
Flowchart and study design of our study. The primary analysis divided the cohort into COVID-19 and matched control groups to assess the presence (primary outcome) and severity (the other primary outcome) of aspergillosis. This study used data from the National Health Insurance Service-National Health Information Database (NHIS-NHID) (NHIS-2022-1-623). The NHIS-NHID database encompasses detailed records of approximately 580,000 confirmed SARS-CoV-2 cases and about 7.9 million individuals in the control group, covering the period from 2020 to 2021. This dataset integrates demographic information, comorbidity profiles, and medical histories collected between 2015 and 2019. The COVID-19 (n = 557,680) and control (n = 557,680) groups were well-matched in terms of demographics, such as sex, age, economic status, and residential area, as well as pre-existing conditions, such as HTN, DM, CKD, CHF, cardiovascular disease (CVD), cancer, MI, asthma, and COPD (all SMDs < 0.1).

Comorbidities were defined using data from the previous 5 years. The comorbidities identified using the ICD-10 codes included angina (I20), cancer (any C code), congestive heart failure (CHF; I43 or I50), chronic kidney disease (CKD; N18 or N19), myocardial infarction (MI; I21 or I22), diabetes mellitus (DM; E10–14), hypertension (HTN; I10–13, I15), asthma (J43 or J44), chronic obstructive pulmonary disease (COPD; J45–46), and chronic liver disease (K74, K703, or B18). Patients with at least one diagnosis within the last 5 years were included in the study. Aspergillosis was identified using ICD-10 codes B44 and B49, with invasive fungal aspergillosis classified under B440 and non-invasive aspergillosis under B441–B449 or B49.

### Statistical Analysis

Descriptive statistics are expressed as frequencies (percentages) for categorical data and means ± standard deviations for continuous data. The standard mean difference (SMD) was used to quantify the differences between the groups, with an SMD < 0.1 indicating a balanced distribution. The COVID-19 and matched control groups were analyzed after 1:1 PS matching to ensure balanced baseline characteristics. Hazard ratios (HRs) were calculated using the Cox regression model. Unadjusted HRs were based on the Kaplan–Meier formula without considering other variables, whereas adjusted HRs accounted for all covariates. Statistical significance was determined using two-sided p-values < 0.05 and 95% confidence intervals (CIs). Analyses were performed using SAS 9.4 and R 4.0.3 (R Foundation for Statistical Computing, Vienna, Austria).

### Transcriptome Analysis

We collected a transcriptome dataset of SARS-Cov-2 infected cell lines from the NCBI Gene Expression Omnibus (GEO) database (https://www.ncbi.nlm.nih.gov/geo/). The datasets included the transcriptome profiles of human lung adenocarcinoma (Calu-3), normal human bronchial epithelial (NHBE) (GSE147507), and human airway epithelial (HAE) cells (GSE153970). The transcriptomes of blood samples from patients with COVID-19 were obtained from EBI ArrayExpress (https://www.ebi.ac.uk/biostudies/arrayexpress) (E-MTAB-10926).

Differentially expressed genes (DEGs) were identified using DESeq2^10^ (FC > 1.5 for cell lines, FC > 2 and FDR < 0.05 for blood samples). Pathway enrichment was calculated as an enrichment factor (EF).

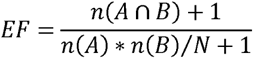

where A is the number of DEGs, B is the number of pathway genes, and N is the total number of genes. Pathway information and protein-protein interaction (PPI) networks were obtained from MSigDB (v2023.2^11^) and STRING (v12.0^12^), respectively.

### Ethics Approval

The study protocol was approved by the Institutional Review Board of Jeonbuk National University Hospital (No. 2024-XX-XXX). The requirement for informed consent was waived because all patient records were anonymized before use.

## Results

### Baseline Characteristics

The baseline characteristics of the study population are shown in Supplementary Table 1. The COVID-19 (n = 557,680) and control (n = 557,680) groups were well matched in terms of demographics such as sex, age, economic status, and residential area, as well as pre-existing conditions such as HTN, DM, CKD, CHF, cardiovascular disease (CVD), cancer, MI, asthma, and COPD (all SMDs < 0.1, Figure 1).

### Incidence and Risk of Aspergillosis in Patients with COVID-19

The incidence and risk of new-onset aspergillosis were compared between COVID-19 and PS-matched control cohorts. New-onset aspergillosis occurred in 0.06% of the COVID-19 group (353 of 557,680) and 0·02% of the control group (94 of 557,680), with incidences of 1.99 and 0.52 per 1000 person-years, respectively (Supplementary Table 2). A significant 3.81-fold difference was observed in the incidence of aspergillosis between the COVID-19 and the matched control groups (HR, 3.81; 95% CI: 3·03–4·78; p < 0.001; Figure 2A). Subgroup analysis showed that the risk of COVID-19-associated respiratory invasive aspergillosis (CARIA) was 27.58 times higher (95% CI: 12.21–62.30) and that for CARNIA was 2.18 times higher (95% CI: 1.69–2.80) in the COVID-19 group compared to the control group (Figures 2B and 2C, Supplementary Table 2). Notably, the risk of aspergillosis was 1.95 times higher (95% CI: 1.51–2.51) in patients with COVID-19 not receiving systemic steroids and 15·68 times higher (95% CI: 12.21–20.14) in patients receiving systemic steroids (Figure 2A, Supplementary Figure 1, Supplementary Table 2). Furthermore, the risk of CARIA was 9.27 times higher (95% CI: 3.96–21.70) in patients with COVID-19 not receiving systemic steroids and 124.89 times higher (95% CI: 54.86–284.34) in patients receiving systemic steroids (Figure 2B, Supplementary Figure 2, Supplementary Table 2). The risk of CARNIA was 1.44 times higher (95% CI: 1.09–1.91) in patients not receiving systemic steroids and 7.35 times higher (95% CI: 5·39–10·02) in patients receiving systemic steroids (Figure 2C, Supplementary Figure 3, Supplementary Table 2). The risk of aspergillosis was higher in males than in females (HR: 1.34, 95% CI: 1.09–1.66), primarily for invasive aspergillosis (HR: 1.80, 95% CI: 1.30–2.48). Younger age was associated with a lower risk of aspergillosis (HR: 0.58, 95% CI: 0.43–0.80), particularly for invasive aspergillosis (HR: 0.18, 95% CI: 0.07–0.44). Individuals aged ≥ 65 years were associated with a higher risk of aspergillosis (HR: 1.81, 95% CI: 1.42–2.31) for both the invasive (HR: 2.17, 95% CI: 1.47–3.20) and non-invasive aspergillosis (HR: 1.59, 95% CI: 1.16–2.18). The presence of comorbidities such as DM, CKD, CHF, cancer, and COPD increased the risk of aspergillosis by 2.69 (95% CI: 2.14–3.38), 1.94 (95% CI: 1.38–2.72), 1.68 (95% CI: 1.32–2.14), 1·34 (95% CI: 1.09–1.66), and 2.36 (95% CI: 1.82–3.07) times, respectively. This trend was consistent for both the CARIA and CARNIA cohorts, except for cancer in CARIA (p = 0.711) and CHF in CARNIA (p = 0.191) (Figures 2A-C, Supplementary Table 2).

**Figure. 2.**
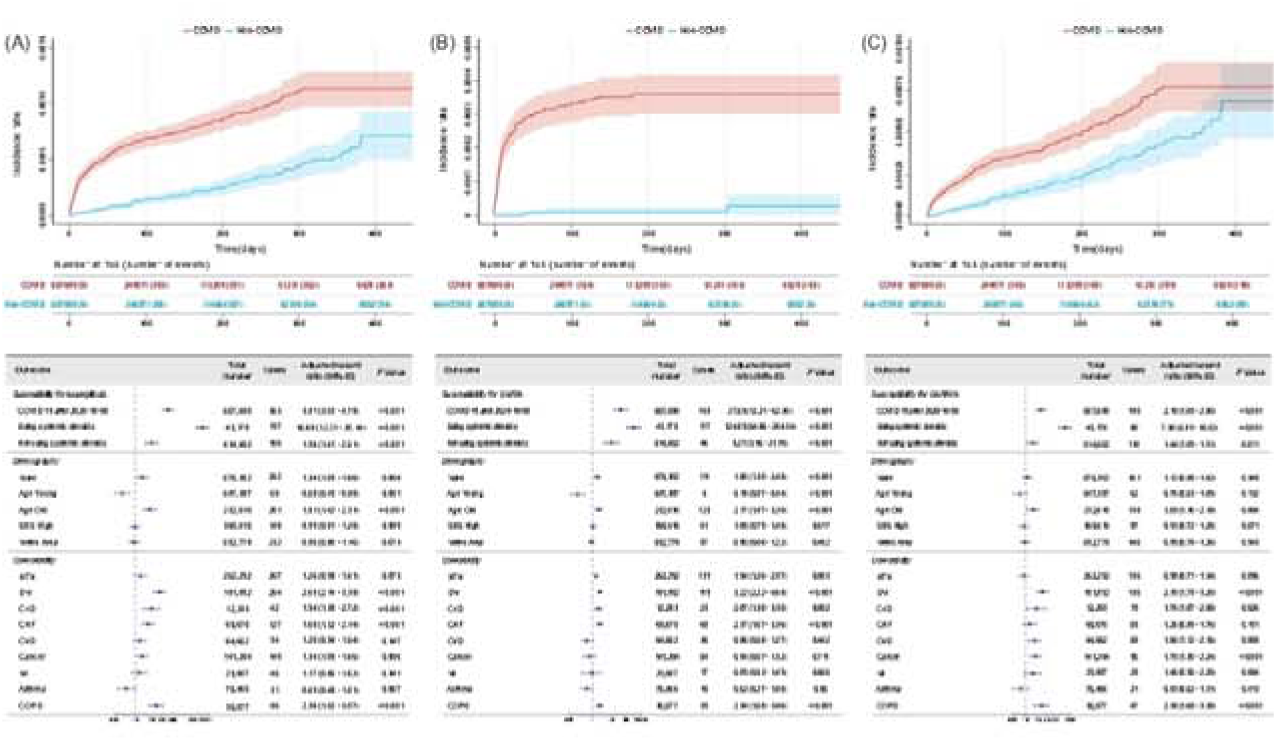
The impact of COVID-19 on the incidence of aspergillosis. (A) Cumulative incidence rate for the aspergillosis susceptibility in the COVID and control (non-COVID) groups; forest plot of the adjusted hazard ratio for each factor: prior COVID-19 infection, use of systemic steroids, demography (male, old age ≥ 65 years), socioeconomic status, comorbidities. (B) Cumulative incidence rate for CARIA susceptibility in the COVID and control (non-COVID) groups; forest plot of the adjusted hazard ratio for each factor: prior COVID-19 infection, use of systemic steroids, demography (male, old age ≥ 65 years), socioeconomic status, comorbidities. (C) Cumulative incidence rate for CARNIA susceptibility in the COVID and control (non-COVID) groups; forest plot of the adjusted hazard ratio for each factor: prior COVID-19 infection, use of systemic steroids, demography (male, old age ≥ 65 years), socioeconomic status, comorbidities.

Economic status and residential area were not significantly associated with aspergillosis risk (HR: 0.99, 95% CI: 0.81–1.20 and HR: 0.96, 95% CI: 0.80–1.16, respectively). Similarly, other comorbidities such as HTN, CVD, MI, and asthma were not significantly associated with the risk of aspergillosis (HR: 1.26, 95% CI: 0.98–1.61; HR: 1.20, 95% CI: 0.94–1.54; HR: 1.17, 95% CI: 0.85–1.62; and HR: 0.69, 95% CI: 0.48–1.01, respectively) (Figure 2A-C, Supplementary Table 2).

### Incidence and Risk of Severe COVID-19 in Patients with Aspergillosis

The incidence and risk of severe COVID-19 in patients with aspergillosis were compared between COVID-19 and PS-matched control cohorts. Severe clinical outcomes occurred in 52.9% of patients with aspergillosis in the COVID-19 group (108 204) and 3.96% of those in the matched control group (22,055 of 557,476), with an incidence of 3254.89 and 165.03 per 1000 person-years, respectively (Figures 3A and 3D, Supplementary Table 3). Patients with aspergillosis had a 1.67-fold higher risk (95% CI: 1.38–2.02) of severe COVID-19 outcomes than those without. More importantly, this was consistent for both CARIA (HR: 1.85, 95% CI: 1.44–2.38) and CARNIA (HR: 1.46, 95% CI: 1.09–1.96) (Figure 3B-D, Supplementary Table 3).

**Figure. 3.**
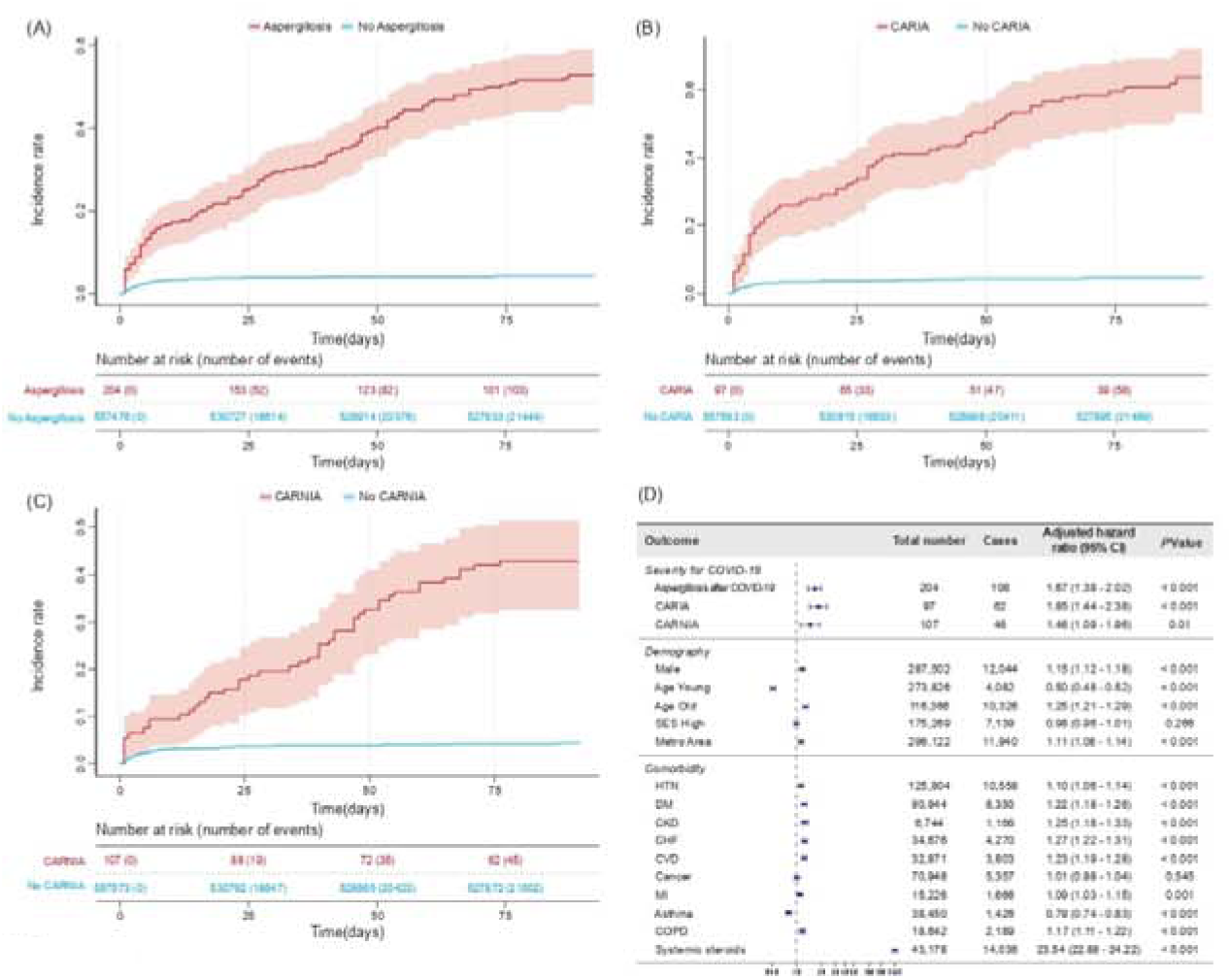
The impact of aspergillosis on COVID-19 severity. (A) Cumulative incidence rate for the severity after COVID-19 in the aspergillosis and control (no aspergillosis) groups. (B) Cumulative incidence rate for the severity after COVID-19 in the CARIA and control (no CARIA) groups. (C) Cumulative incidence rate for the severity after COVID-19 in the CARNIA and control (no CARNIA) groups. (D) Forest plot of the adjusted hazard ratio for each factor: aspergillosis, CARIA, CARNIA group at risk for severe COVID-19, demography (male, old age ≥ 65 years), socioeconomic status, comorbidities.

This trend was more pronounced in males than in females (HR: 1.15, 95% CI: 1.12–1.18) for both CARIA and CARNIA. Younger age was associated with a lower risk of severe COVID-19 (HR: 0.50, 95% CI: 0.48–0.52), while older age was associated with a higher risk (HR: 1.25, 95% CI: 1.21–1.29); these findings were consistent for both CARIA and CARNIA. Urban residents had a 1.11-fold higher risk of severe COVID-19 outcomes compared to other residents (95% CI: 1.08–1.14) (Figure 3D, Supplementary Table 3).

Comorbidities such as HTN, DM, CKD, CHF, CVD, MI, and COPD were associated with an increased risk of severe COVID-19 outcomes by 1.10 (95% CI: 1.06–1.14), 1.22 (95% CI: 1.18–1.26), 1.25 (95% CI: 1.18–1.33), 1.27 (95% CI: 1.22–1.31), 1.23 (95% CI: 1.19–1.28), 1.09 (95% CI: 1.03–1.15), and 1.17 (95% CI: 1.11–1.22) times, respectively. These associations were consistent in both CARIA and CARNIA (Figure 3D, Supplementary Table 3). Notably, the risk of severe COVID-19 outcomes was 23.54 times higher (95% CI: 22.88– 24.22) in patients receiving systemic steroids than in those that did not (Figure 3D, Supplementary Table 3).

### Transcriptomic Changes after SARS-CoV-2 Infection and Steroid Treatment

To explore the molecular pathobiological context of our epidemiological findings, we analyzed the transcriptome profiles of SARS-CoV-2-infected epithelial cell lines and COVID-19 patient blood samples (Figure 4A), focusing on inflammatory pathways associated with fungal infections.

**Figure. 4.**
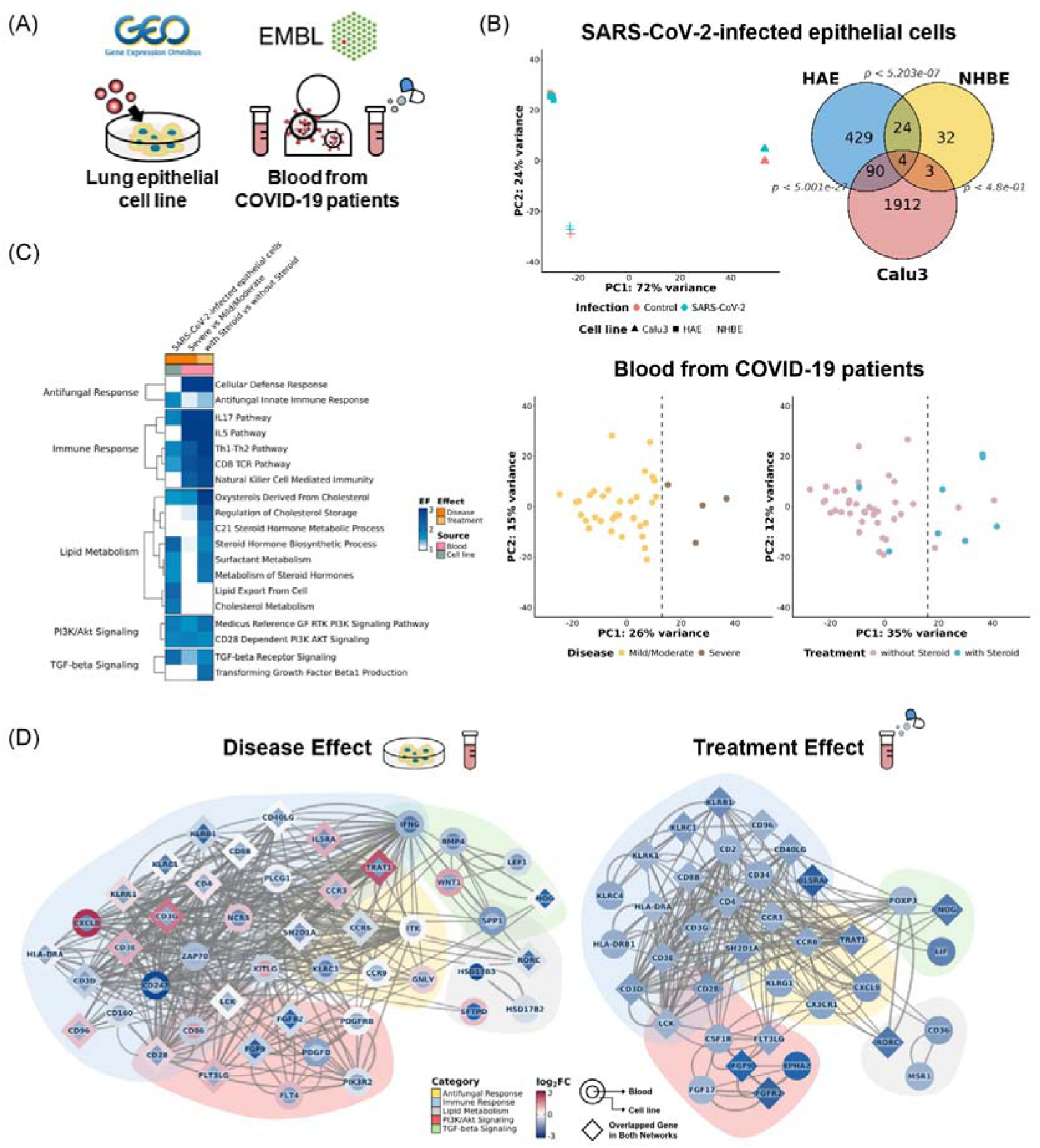
The antifungal responses are downregulated at the transcriptional level. (A) A schematic representation of the public transcriptome dataset, which was obtained from two different sources (cell lines and blood) and categorized based on study design: SARS-CoV-2 infection and steroid treatment after infection. (B) Principal component analysis (PCA) plot of the study design. Genes differentially expressed in cell lines overlapped with each other. (C) Significantly enriched pathways with downregulated DEG sets were categorized into five groups: antifungal response, immune response, lipid metabolism, PI3K/Akt signaling, and TGF-β signaling. (D) Protein-protein interaction (PPI) involved in five key groups of pathways. The interactions were obtained from STRING and filtered using a confidence score cutoff (> 0.7). Inner nodes are colored by the fold change of gene expression (log2FC) in immunological aspects, and the outer color represents mean log2FC in structural aspects. The weight of edge indicates the interaction strength between proteins. Overlapped proteins between disease and treatment effect are denoted by diamond.

First, we compared the DEGs of the three infected epithelial cell lines and identified 121 down-DEGs common to at least two cell lines. For patient blood samples, we observed a distinct grouping of expression patterns according to disease severity and steroid treatment/non-treatment (Figure 4B). Antifungal and immune-related pathways were consistently suppressed by SARS-CoV-2 infection, disease progression, and steroid treatment (Figure 4C). SARS-CoV-2 attenuated T helper 2 (Th2)-type responses (e.g., interleukin-5), which drive eosinophil-mediated clearance^13^ in blood cells, and downstream interleukin-17 signaling that reduces *Af* burden^14^ in both epithelial and immune cells. NK cell dysfunction is also impaired in blood cells. It may contribute to reduced cytotoxic activity by perforin, compromising antifungal defense^15^. Furthermore, lipid metabolic pathways (e.g., cholesterol, surfactants), essential for host’s bronchial antifungal immunity^16^, phagocytosis^17^, and enhancing antifungal drug activity^18^, were downregulated in SARS-CoV-2-infected epithelial cells. Lastly, phosphatidylinositol 3-kinase (PI3K) signaling pathways, involving in the recognition of *Af* conidia by neutrophils via integrin CD11b/CD18^19^, were also significantly downregulated in both epithelial and immune cells in our data.

We then investigated the interactions between the downregulated pathway genes using the STRING protein-protein network (Figure 4D).^12^ These genes formed highly connected networks in both disease and treatment cases, contributing to impaired antifungal responses. (p < 0.000175 and p < 0.000167, respectively, estimated by sampling 1,000,000 random subnetworks of the same number of nodes). Interferon-gamma (IFN-γ) signaling, including CD4, IFNG, and CXCL9, emerged as significant nodes in our network. CD4 is the top connected node in both networks, which activates T cells by toll-like receptor (TLR)- dependent IFN-γ production^20^. IFN-γ shows a synergistic effect with antifungal drugs (e.g., amphotericin B)^21^ and induces CXCL9, a CXC chemokine^22^ that enhances NADPH oxidase activity through plasmacytoid dendritic cells (DCs)^23^. Specifically, CD4 is consistently downregulated and the most connected node across all networks, while IFNG and CXCL9 are prominently present only in disease and treatment networks, respectively. Additionally, CD3E and CCR6 are notably connected within both networks and CX3CR1 is observed specifically in the treatment network. CD3E promotes IL-10-mediated repair in *Af* keratitis^24^. CX3CR1, which is stimulated by CD3^25^, is negatively correlated with the risk of invasive aspergillosis^26^. CCR6 mediates the migration of myeloid dendritic cells (DCs) during *Af* infection^27^.

## Discussion

This is the first report to conduct a comprehensive analysis of a substantial nationwide cohort of 8.5 million clinical registries with CARIA and CARNIA in the respiratory system. The incidence of both forms of aspergillosis has increased in patients with a SARS-CoV-2 infection. Notably, it was corroborated that the severity including mortality of COVID-19 increased remarkably not only in patients with CARIA, but also in those with CARNIA.

Recent studies have reported an increased incidence of invasive aspergillosis in patients with COVID-19 in the ICU, and various underlying pathophysiological mechanisms have been suggested.^3,4,28^ Our data validated the increased incidence of CARIA. SARS-CoV-2 infection itself is a cardinal risk factor for developing CARIA, and as previously known, comorbidities also contribute to the increased incidence.^28^ Interestingly, the incidence of CARNIA significantly increased in patients with COVID-19. Of particular importance is the substantially increased incidence of CARNIA in individuals with comorbidities, such as DM and COPD. Moreover, although little is known about CARNIA, we have recently demonstrated through preclinical models of *Af*-induced severe allergic lung inflammation, one of the clinical spectrums of non-invasive aspergillosis, that a COVID-19-induced dysregulation of innate immune response is closely involved in the clinical course of the disease.^8^ Thus, immunological changes owing to viral infections in the micromilieu surrounding the respiratory epithelium could prime the host respiratory tract for the development and aggravation of subsequent aspergillosis, including both invasive and non-invasive forms.

Furthermore, CAPA, a term tending towards invasive aspergillosis, has been reported to increase the mortality of patients with COVID-19, and in accordance with previous studies, the severity was 1.85 times higher in CARIA.^3,4^ Notably, the severity was also 1.46 times higher even in non-invasive aspergillosis, which was corroborated for the first time. The increased severity of COVID-19 caused by CARNIA was also substantiated for the first time. With respect to CARIA, invasive aspergillosis is reported to increase the severity of COVID-19 in patients through a cytokine storm.^28,29,30^ More importantly, despite the limited knowledge on non-invasive aspergillosis, we have previously demonstrated with animal models and large clinical data that dysregulated immune response associated with underlying non-invasive pulmonary aspergillosis contributes to the increased mortality of patients with COVID-19. CARNIA may activate several crucial innate immune components in the cells, such as NLRP3 inflammasome, thereby inducing pro-inflammatory cytokines (interleukin [IL]-1, IL-6, IL-17, and tumor necrosis factor) that increase the severity of COVID-19.^8^

In the era of COVID-19, comorbidities, such as DM, COPD, HTN and CHF, have been highlighted as key drivers of increased COVID-19 severity.^31^ Furthermore, our group recently reported the impact of prior respiratory syncytial virus infection within 3 years on the severity of COVID-19.^9^ In light of this, host-related medical factors, such as comorbidities and previous medical history, may play pivotal roles in increasing the severity of COVID-19 not only in patients with invasive aspergillosis but also in those with non-invasive aspergillosis. As expected, comorbidities including CKD, CHF, and CVD worsened COVID-19 in patients with subsequent aspergillosis. Additionally, our study confirmed that the increased severity of CARIA or CARNIA could be attributed to COVID-19-targeted treatment. This is consistent with the accumulating evidence that a prolonged treatment course with systemic steroid therapy in hospitalized patients with COVID-19 is correlated with higher mortality and poor outcomes.^32^ However, further studies are required to delineate clear mechanisms.

Importantly, the findings from our epidemiological data were validated at the molecular level using a public dataset, particularly for human-derived systems, including human cell lines and patient blood samples. Notably, SARS-CoV-2 infection weakened the antifungal defense pathway. Marked downregulation of immune response, such as T cell, interleukin, and NK cell mediated pathway were significantly diminished. Furthermore, the suppression of lipid metabolism and PI3K/Akt signaling pathways, both of which are known to play pivotal roles in fungal defense, was evident. Through supplementary analysis using the protein–protein interaction network, we further identified highly interconnected nodes associated with the observed impairment in antifungal immunity, particularly involving genes related to IFN-γ signaling. Notably, the CD4, IFNG, CXCL9, CD3E, CCR6, and CX3CR1 emerged as central components within this network. These genes have previously been recognized as key regulators of antifungal defense, thereby reinforcing the validity of our epidemiological findings^20–27^. These nodes represent promising targets for preclinical research that focuses on patients vulnerable to secondary fungal infections following viral illnesses. Targeting these molecules may pave the way for translational strategies aimed at developing novel pathogen-specific therapeutics.

## Conclusion

In this study, utilising hitherto the largest cohort of 8.5 million clinical registries, we demonstrated that SARS-CoV-2 increases the incidence of both invasive and non-invasive forms of aspergillosis in the respiratory system. Notably, in addition to the well-known impact of CAPA on the severity of COVID-19, our data demonstrate, for the first time, that CARNIA significantly increases the severity of COVID-19. Moreover, systemic steroids have deleterious effects on the severity of COVID-19 in patients with subsequent aspergillosis. Our results highlight the therapeutic potential of antifungal agents for both invasive and non-invasive aspergillosis to improve the outcomes of patients infected with respiratory viruses, although results from well-designed clinical trials are warranted in the near future. Additionally, clinicians responsible for the care of patients with respiratory viral infections must maintain a high level of surveillance and be aware of the impact of excessive medical treatments such as systemic steroids on patients.

**Table 1.**
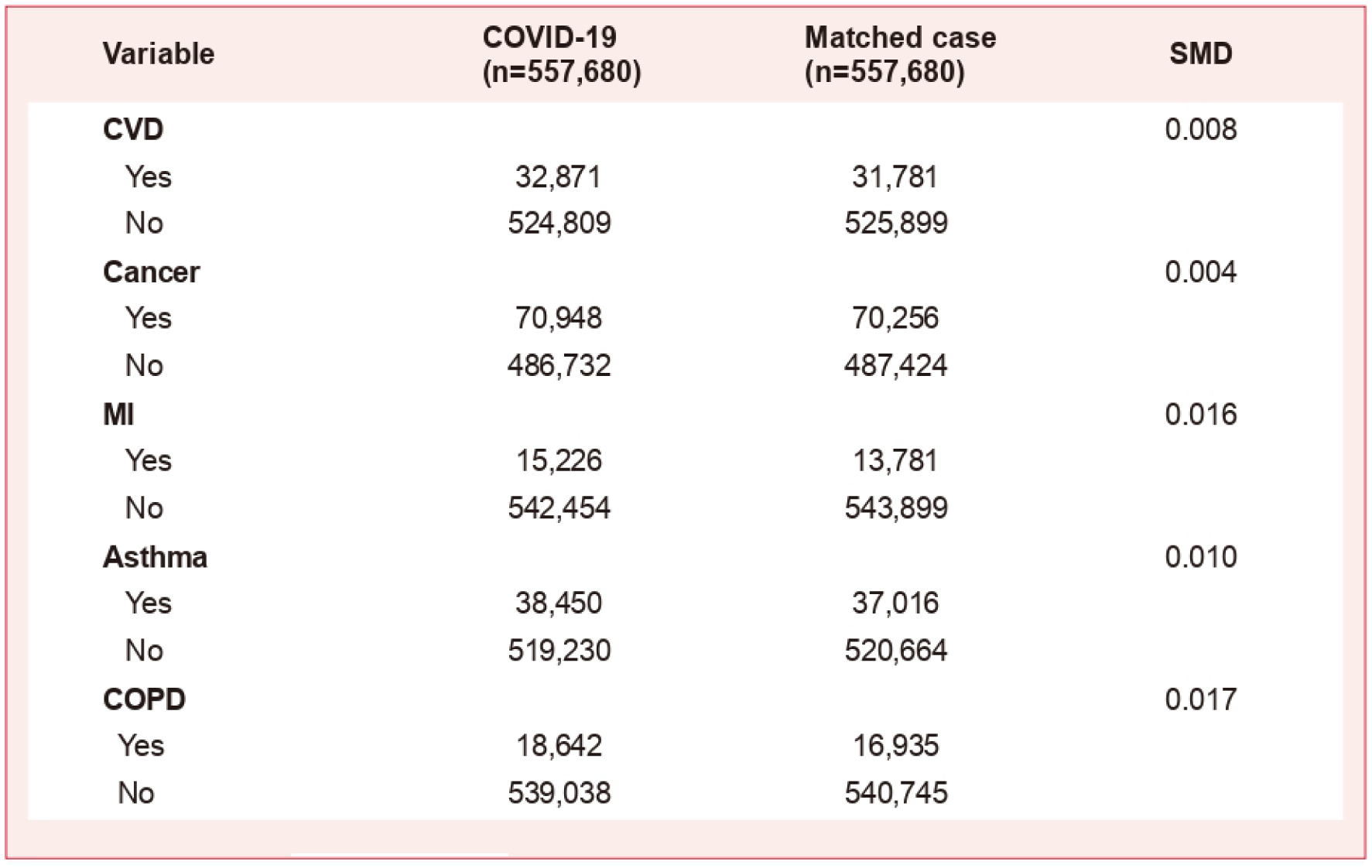
the SMDs between COVID-19 and matched cases after the propensity score matching. SMD, the standardized mean difference; HTN, hypertension; DM, diabetes mellitus; CKD, chronic kidney disease; CHF, congestive heart failure; CVD, cardiovascular disease; MI, myocardial infarction; COPD, chronic obstructive pulmonary disease;

**Table 2.** Incidence per 1,000 person-years and hazard ratios for fungal infection including invasive and non-invasive fungal infection. HTN, hypertension; DM, diabetes mellitus; CKD, chronic kidney disease; CHF, congestive heart failure; CVD, cardiovascular disease; MI, myocardial infarction; COPD, chronic obstructive pulmonary disease

| Variable | Total | Cases | Incidence<br>per 1,000<br>person-years | Unadjusted<br>hazard ratio | Adjusted<br>hazard ratio | P Value |
| --- | --- | --- | --- | --- | --- | --- |
| **Sub : CARNIA** |  |  |  |  |  |  |
| **HTN** |  |  |  |  |  |  |
| Yes | 252,292 | 136 | 1.68 | 3.28 (2.59-4.15) | 0.98 (0.71-1.34) | 0.896 |
| No | 863,068 | 142 | 0.51 | 1 | 1 |  |
| **DM** |  |  |  |  |  |  |
| Yes | 181,892 | 135 | 2.31 | 4.83 (3.82-6.12) | 2.39 (1.78-3.20) | < 0.001 |
| No | 933,468 | 143 | 0.48 | 1 | 1 |  |
| **CKD** |  |  |  |  |  |  |
| Yes | 12,283 | 19 | 5.16 | 6.93 (4.35-11.04) | 1.76 (1.07-2.88) | 0.026 |
| No | 1,103,077 | 259 | 0.73 | 1 | 1 |  |
| **CHF** |  |  |  |  |  |  |
| Yes | 68,670 | 59 | 2.63 | 4.03 (3.02-5.38) | 1.25 (0.89-1.76) | 0.191 |
| No | 1,046,690 | 219 | 0.65 | 1 | 1 |  |
| **CVD** |  |  |  |  |  |  |
| Yes | 64,652 | 58 | 2.86 | 4.35 (3.26-5.82) | 1.56 (1.12-2.15) | 0.008 |
| No | 1,050,708 | 220 | 0.65 | 1 | 1 |  |
| **Cancer** |  |  |  |  |  |  |
| Yes | 141,204 | 95 | 2.05 | 3.50 (2.73-4.49) | 1.70 (1.30-2.24) | < 0.001 |
| No | 974,156 | 183 | 0.59 | 1 | 1 |  |
| **MI** |  |  |  |  |  |  |
| Yes | 29,007 | 28 | 2.91 | 4.07 (2.75-6.01) | 1.45 (0.95-2.20) | 0.084 |
| No | 1,086,353 | 250 | 0.72 | 1 | 1 |  |
| **Asthma** |  |  |  |  |  |  |
| Yes | 75,466 | 21 | 1.00 | 1.26 (0.81-1.97) | 0.83 (0.52-1.31) | 0.413 |
| No | 1,039,894 | 257 | 0.76 | 1 | 1 |  |
| **COPD** |  |  |  |  |  |  |
| Yes | 35,577 | 47 | 4.13 | 6.16 (4.50-8.44) | 2.38 (1.68-3.38) | < 0.001 |
| No | 1,079,783 | 231 | 0.67 | 1 | 1 |  |

**Table 3.** Incidence per 1,000 person-years and hazard ratios for severity of COVID-19. HTN, hypertension; DM, diabetes mellitus; CKD, chronic kidney disease; CHF, congestive heart failure; CVD, cardiovascular disease; MI, myocardial infarction; COPD, chronic obstructive pulmonary disease

| Variable | Total | Cases | Incidence<br>per 1,000<br>person-years | Unadjusted<br>hazard ratio | Adjusted<br>hazard ratio | P Value |
| --- | --- | --- | --- | --- | --- | --- |
| **CKD** |  |  |  |  |  |  |
| Yes | 6,744 | 1,166 | 845.88 | 4.96 (4.68-5.26) | 1.25 (1.18-1.33) | < 0.001 |
| No | 550,936 | 20,997 | 158.71 | 1 | 1 |  |
| **CHF** |  |  |  |  |  |  |
| Yes | 34,676 | 4,270 | 573.29 | 3.87 (3.75-4.00) | 1.27 (1.22-1.31) | < 0.001 |
| No | 523,004 | 17,893 | 141.75 | 1 | 1 |  |
| **CVD** |  |  |  |  |  |  |
| Yes | 32,871 | 3,803 | 530.30 | 3.51 (3.39-3.64) | 1.23 (1.19-1.28) | < 0.001 |
| No | 524,809 | 18,360 | 145.13 | 1 | 1 |  |
| **Cancer** |  |  |  |  |  |  |
| Yes | 70,948 | 5,357 | 327.82 | 2.25 (2.18-2.32) | 1.01 (0.98-1.04) | 0.545 |
| No | 486,732 | 16,806 | 143.23 | 1 | 1 |  |
| **MI** |  |  |  |  |  |  |
| Yes | 15,226 | 1,666 | 495.44 | 3.04 (2.89-3.20) | 1.09 (1.03-1.15) | 0.001 |
| No | 542,454 | 20,497 | 157.29 | 1 | 1 |  |
| **Asthma** |  |  |  |  |  |  |
| Yes | 38,450 | 1,426 | 154.44 | 0.93 (0.88-0.98) | 0.79 (0.74-0.83) | < 0.001 |
| No | 519,230 | 20,737 | 166.64 | 1 | 1 |  |
| **COPD** |  |  |  |  |  |  |
| Yes | 18,642 | 2,189 | 541.35 | 3.38 (3.23-3.53) | 1.17 (1.11-1.22) | < 0.001 |
| No | 539,038 | 19,974 | 154.08 | 1 | 1 |  |
| **Systemic steroids** |  |  |  |  |  |  |
| Yes | 43,178 | 14,036 | 2351.31 | 38.32 (37.30-39.37) | 23.54 (22.88-24.22) | < 0.001 |
| No | 514,502 | 8,127 | 63.64 | 1 | 1 |  |

## Data Availability

The data supporting the findings of this study are available from the National Health Insurance Service-National Health Information Database (NHIS-NHID) (NHIS-2022-1-623) of the Republic of Korea. These requests should be addressed to Jong Seung Kim.

## Contributors

JHP, YCL, JK, WK and JSJ interpreted the data and wrote the manuscript. JSK, JWY, and JK conducted statistical planning and data analysis. JSJ initially conceived the study. JSJ, JSK, WK and YCL designed and supervised the project. All the authors critically reviewed and approved the final version of the manuscript.

## Acknowledgments

This work was supported by the National Research Foundation of Korea (NRF) grant funded by the Korea government (MSIT) (No. RS-2024-00356349; JSJ, No. RS-2022-NR069855; WK, No. RS-2025-00514175; JSK) and Special Operating Subsidy of Jeonbuk National University Industrial Cooperation Foundation. This research was supported by a grant of the Korea Health Technology R&D Project through the Korea Health Industry Development Institute (KHIDI), funded by the Ministry of Health & Welfare, Republic of Korea (grant number: RS-2024-00440408; JSJ). This paper was supported by Fund of Biomedical Research Institute, Jeonbuk National University Hospital.

---

**Supplementary Figure. 1.**
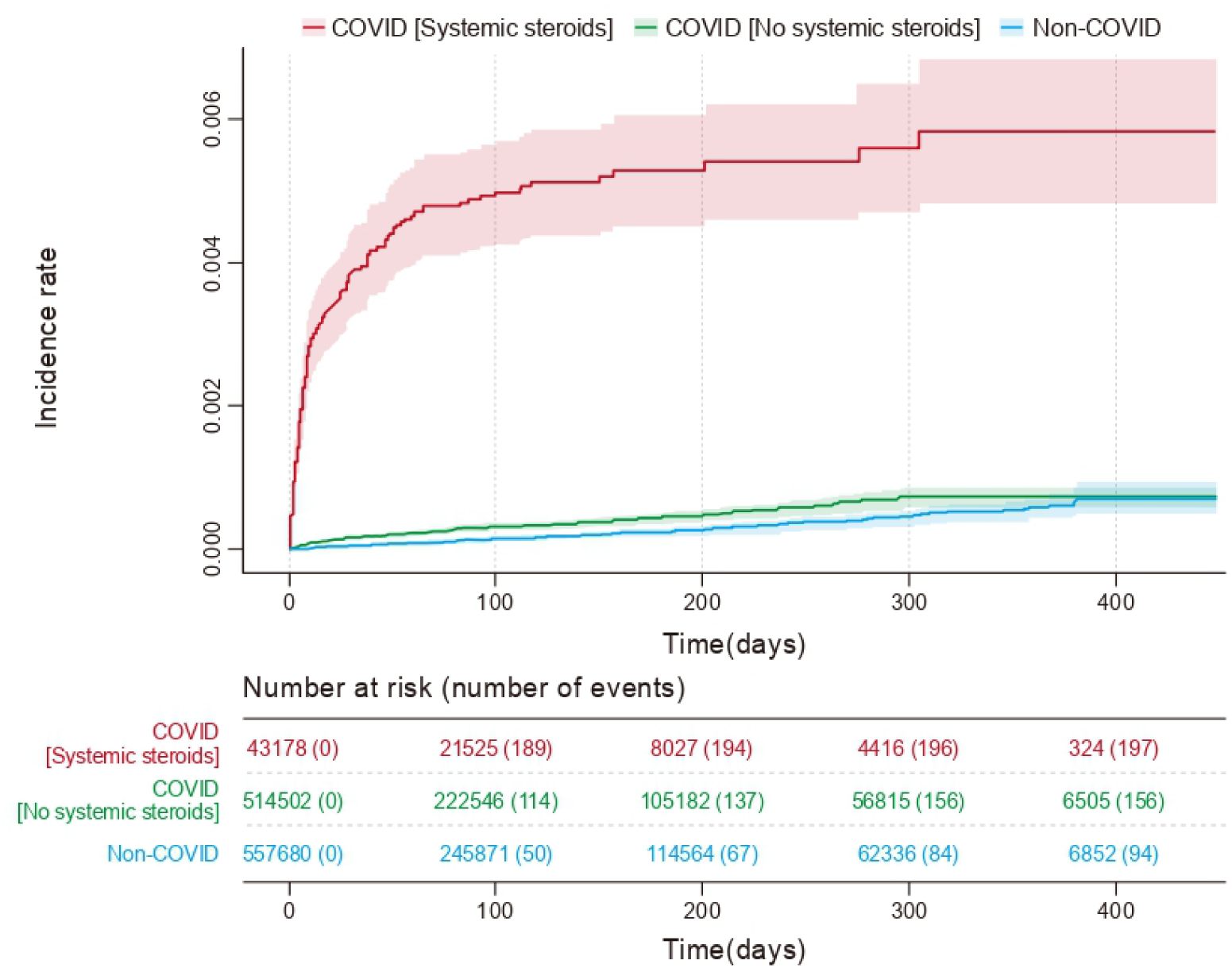

**Supplementary Figure. 2.**
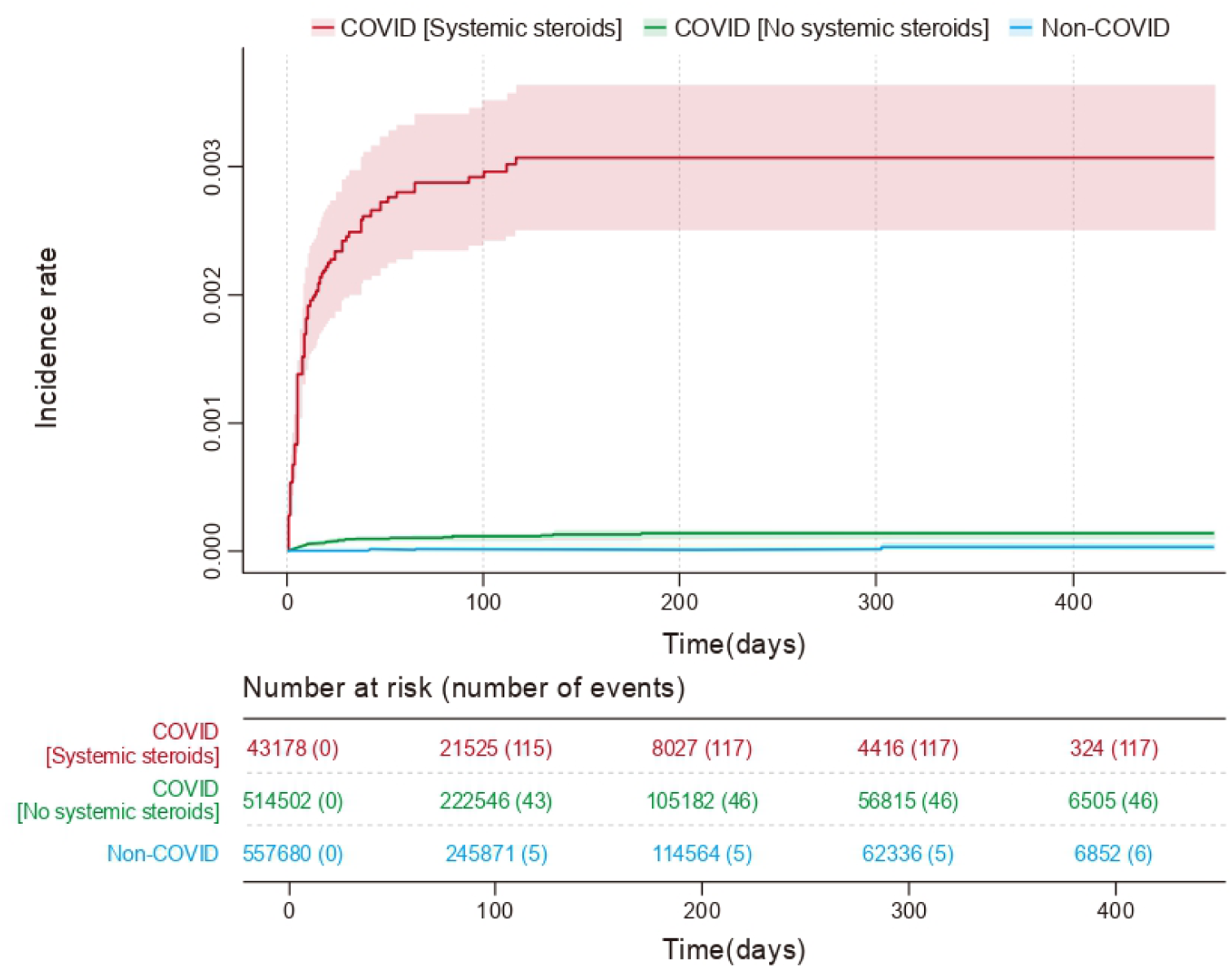

**Supplementary Figure. 3.**
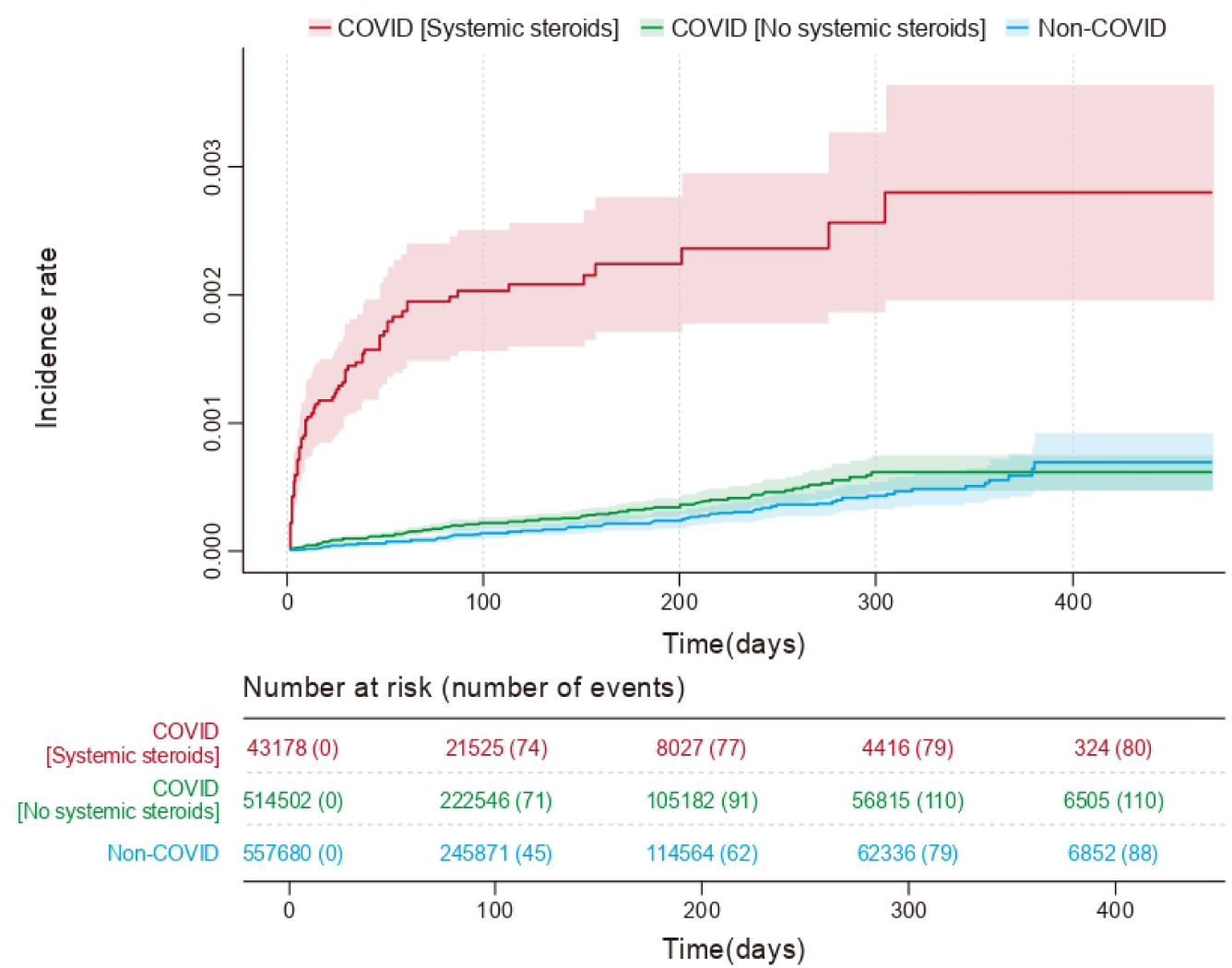

**Supplementary Table. 1.**

| Variable | COVID-19<br>(n=557,680) | Matched case<br>(n=557,680) | SMD |
| --- | --- | --- | --- |
| **Sex** |  |  | 0.001 |
| Male | 287,502 | 287,800 |  |
| Female | 270,178 | 269,880 |  |
| **Age** |  |  | 0.002 |
| Young | 273,826 | 273,561 |  |
| Middle | 167,488 | 167,875 |  |
| Old | 116,366 | 116,244 |  |
| **Economic status** |  |  | < 0.001 |
| High | 175,269 | 175,346 |  |
| Low | 382,411 | 382,334 |  |
| **Region** |  |  | 0.002 |
| Metro | 296,122 | 296,656 |  |
| Rural | 261,558 | 261,024 |  |
| **HTN** |  |  | 0.003 |
| Yes | 125,804 | 126,488 |  |
| No | 431,876 | 431,192 |  |
| **DM** |  |  | < 0.001 |
| Yes | 90,944 | 90,948 |  |
| No | 466,736 | 466,732 |  |
| **CKD** |  |  | 0.021 |
| Yes | 6,744 | 5,539 |  |
| No | 550,936 | 552,141 |  |
| **CHF** |  |  | 0.005 |
| Yes | 34,676 | 33,994 |  |
| No | 523,004 | 523,686 |  |

**Supplementary Table. 2.**

| Variable | Total | Cases | Incidence<br>per 1,000<br>person-years | Unadjusted<br>hazard ratio | Adjusted<br>hazard ratio | P Value |
| --- | --- | --- | --- | --- | --- | --- |
| **COVID-19-associated Aspergillosis** |  |  |  |  |  |  |
| **Total** | 1,115,360 | 447 |  |  |  |  |
| **COVID-19** |  |  |  |  |  |  |
| Yes | 557,680 | 353 | 1.99 | 3.78 (3.01-4.74) | 3.81 (3.03-4.78) | < 0.001 |
| Systemic steroids Yes | 43,178 | 197 | 13.76 | 26.36 (20.61-33.70) | 15.68 (12.21-20.14) | < 0.001 |
| Systemic steroids No | 514,502 | 156 | 0.95 | 1.81 (1.40-2.34) | 1.95 (1.51-2.51) | < 0.001 |
| No | 557,680 | 94 | 0.52 | 1 | 1 |  |
| **Sex** |  |  |  |  |  |  |
| Male | 575,302 | 262 | 1.43 | 1.33 (1.10-1.60) | 1.34 (1.09-1.66) | 0.004 |
| Female | 540,058 | 185 | 1.07 | 1 | 1 |  |
| **Age** |  |  |  |  |  |  |
| Young | 547,387 | 68 | 0.40 | 0.38 (0.28-0.51) | 0.58 (0.43-0.80) | 0.001 |
| Old | 232,610 | 261 | 3.76 | 3.54 (2.85-4.40) | 1.81 (1.42-2.31) | < 0.001 |
| Middle | 335,363 | 118 | 1.00 | 1 | 1 |  |
| **Economic status** |  |  |  |  |  |  |
| High | 350,615 | 148 | 1.33 | 1.09 (0.90-1.33) | 0.99 (0.81-1.20) | 0.889 |
| Low | 764,745 | 299 | 1.22 | 1 | 1 |  |
| **Region** |  |  |  |  |  |  |
| Metro | 592,778 | 232 | 1.25 | 0.98 (0.82-1.18) | 0.96 (0.80-1.16) | 0.673 |
| Rural | 522,582 | 215 | 1.25 | 1 | 1 |  |
| **HTN** |  |  |  |  |  |  |
| Yes | 252,292 | 267 | 3.31 | 5.11 (4.23-6.18) | 1.26 (0.98-1.61) | 0.073 |
| No | 863,068 | 180 | 0.65 | 1 | 1 |  |
| **DM** |  |  |  |  |  |  |
| Yes | 181,892 | 254 | 4.35 | 6.78 (5.62-8.18) | 2.69 (2.14-3.38) | < 0.001 |
| No | 933,468 | 193 | 0.65 | 1 | 1 |  |
| **CKD** |  |  |  |  |  |  |
| Yes | 12,283 | 42 | 11.41 | 9.75 (7.09-13.39) | 1.94 (1.38-2.72) | < 0.001 |
| No | 1,103,077 | 405 | 1.15 | 1 | 1 |  |

Supplementary Table.2
| Variable | Total | Cases | Incidence<br>per 1,000<br>person-years | Unadjusted<br>hazard ratio | Adjusted<br>hazard ratio | P Value |
| --- | --- | --- | --- | --- | --- | --- |
| **COVID-19-associated Aspergillosis** |  |  |  |  |  |  |
| **CHF** |  |  |  |  |  |  |
| Yes | 68,670 | 127 | 5.67 | 5.99 (4.88-7.36) | 1.68 (1.32-2.14) | < 0.001 |
| No | 1,046,690 | 320 | 0.96 | 1 | 1 |  |
| **CVD** |  |  |  |  |  |  |
| Yes | 64,652 | 94 | 4.64 | 4.41 (3.51-5.54) | 1.20 (0.94-1.54) | 0.147 |
| No | 1,050,708 | 353 | 1.05 | 1 | 1 |  |
| **Cancer** |  |  |  |  |  |  |
| Yes | 141,204 | 149 | 3.21 | 3.41 (2.80-4.15) | 1.34 (1.09-1.66) | 0.006 |
| No | 974,156 | 298 | 0.96 | 1 | 1 |  |
| **MI** |  |  |  |  |  |  |
| Yes | 29,007 | 45 | 4.68 | 4.12 (3.02-5.60) | 1.17 (0.85-1.62) | 0.341 |
| No | 1,086,353 | 402 | 1.16 | 1 | 1 |  |
| **Asthma** |  |  |  |  |  |  |
| Yes | 75,466 | 31 | 1.47 | 1.12 (0.78-1.62) | 0.69 (0.48-1.01) | 0.057 |
| No | 1,039,894 | 416 | 1.24 | 1 | 1 |  |
| **COPD** |  |  |  |  |  |  |
| Yes | 35,577 | 86 | 7.55 | 7.26 (5.73-9.18) | 2.36 (1.82-3.07) | < 0.001 |
| No | 1,079,783 | 361 | 1.04 | 1 | 1 |  |
| **Sub : CARIA** |  |  |  |  |  |  |
| **Total** | 1,115,360 | 169 |  |  |  |  |
| **COVID-19** |  |  |  |  |  |  |
| Yes | 557,680 | 163 | 0.92 | 27.25 (12.07-61.56) | 27.58 (12.21-62.30) | < 0.001 |
| Systemic steroids Yes | 43,178 | 117 | 8.17 | 243.48 (107.19-553.06) | 124.89 (54.86-284.34) | < 0.001 |
| Systemic steroids No | 514,502 | 46 | 0.28 | 8.36 (3.57-19.58) | 9.27 (3.96-21.70) | < 0.001 |
| No | 557,680 | 6 | 0.03 | 1 | 1 |  |
| **Sex** |  |  |  |  |  |  |
| Male | 575,302 | 111 | 0.60 | 1.78 (1.30-2.45) | 1.80 (1.30-2.48) | < 0.001 |
| Female | 540,058 | 58 | 0.33 | 1 | 1 |  |

Supplementary Table.2
| Variable | Total | Cases | Incidence<br>per 1,000<br>person-years | Unadjusted<br>hazard ratio | Adjusted<br>hazard ratio | P Value |
| --- | --- | --- | --- | --- | --- | --- |
| **Sub : CARIA** |  |  |  |  |  |  |
| **Age** |  |  |  |  |  |  |
| Young | 547,387 | 6 | 0.04 | 0.09 (0.04-0.22) | 0.18 (0.07-0.44) | < 0.001 |
| Old | 232,610 | 123 | 1.77 | 4.69 (3.28-6.71) | 2.17 (1.47-3.20) | < 0.001 |
| Middle | 335,363 | 40 | 0.34 | 1 | 1 |  |
| **Economic status** |  |  |  |  |  |  |
| High | 350,615 | 61 | 0.55 | 1.25 (0.91-1.71) | 1.08 (0.79-1.48) | 0.617 |
| Low | 764,745 | 108 | 0.44 | 1 | 1 |  |
| **Region** |  |  |  |  |  |  |
| Metro | 592,778 | 87 | 0.47 | 0.95 (0.70-1.28) | 0.90 (0.66-1.22) | 0.492 |
| Rural | 522,582 | 82 | 0.48 | 1 | 1 |  |
| **HTN** |  |  |  |  |  |  |
| Yes | 252,292 | 131 | 1.62 | 11.98 (8.35-17.20) | 1.94 (1.26-2.97) | 0.003 |
| No | 863,068 | 38 | 0.14 | 1 | 1 |  |
| **DM** |  |  |  |  |  |  |
| Yes | 181,892 | 119 | 2.04 | 12.38 (8.89-17.22) | 3.22 (2.22-4.68) | < 0.001 |
| No | 933,468 | 50 | 0.17 | 1 | 1 |  |
| **CKD** |  |  |  |  |  |  |
| Yes | 12,283 | 23 | 6.25 | 14.68 (9.46-22.79) | 2.07 (1.30-3.30) | 0.002 |
| No | 1,103,077 | 146 | 0.41 | 1 | 1 |  |
| **CHF** |  |  |  |  |  |  |
| Yes | 68,670 | 68 | 3.03 | 10.31 (7.58-14.02) | 2.37 (1.67-3.36) | < 0.001 |
| No | 1,046,690 | 101 | 0.30 | 1 | 1 |  |
| **CVD** |  |  |  |  |  |  |
| Yes | 64,652 | 36 | 1.78 | 4.50 (3.11-6.50) | 0.86 (0.58-1.27) | 0.442 |
| No | 1,050,708 | 133 | 0.39 | 1 | 1 |  |
| **Cancer** |  |  |  |  |  |  |
| Yes | 141,204 | 54 | 1.16 | 3.26 (2.36-4.50) | 0.94 (0.67-1.32) | 0.711 |
| No | 974,156 | 115 | 0.37 | 1 | 1 |  |

Supplementary Table.2
| Variable | Total | Cases | Incidence<br>per 1,000<br>person-years | Unadjusted<br>hazard ratio | Adjusted<br>hazard ratio | P Value |
| --- | --- | --- | --- | --- | --- | --- |
| **Sub : CARIA** |  |  |  |  |  |  |
| **MI** |  |  |  |  |  |  |
| Yes | 29,007 | 17 | 1.77 | 4.19 (2.54-6.92) | 0.89 (0.53-1.49) | 0.655 |
| No | 1,086,353 | 152 | 0.44 | 1 | 1 |  |
| **Asthma** |  |  |  |  |  |  |
| Yes | 75,466 | 10 | 0.48 | 0.91 (0.48-1.72) | 0.52 (0.27-1.00) | 0.050 |
| No | 1,039,894 | 159 | 0.47 | 1 | 1 |  |
| **COPD** |  |  |  |  |  |  |
| Yes | 35,577 | 39 | 3.42 | 9.22 (6.44-13.18) | 2.34 (1.58-3.46) | < 0.001 |
| No | 1,079,783 | 130 | 0.38 | 1 | 1 |  |
| **Sub : CARNIA** |  |  |  |  |  |  |
| **Total** | 1,115,360 | 278 |  |  |  |  |
| **COVID-19** |  |  |  |  |  |  |
| Yes | 557,680 | 190 | 1.07 | 2.17 (1.69-2.80) | 2.18 (1.69-2.80) | < 0.001 |
| Systemic steroids Yes | 43,178 | 80 | 5.59 | 11.48 (8.48-15.53) | 7.35 (5.39-10.02) | < 0.001 |
| Systemic steroids No | 514,502 | 110 | 0.67 | 1.37 (1.03-1.81) | 1.44 (1.09-1.91) | 0.011 |
| No | 557,680 | 88 | 0.49 | 1 | 1 |  |
| **Sex** |  |  |  |  |  |  |
| Male | 575,302 | 151 | 0.82 | 1.12 (0.88-1.42) | 1.12 (0.88-1.42) | 0.349 |
| Female | 540,058 | 127 | 0.73 | 1 | 1 |  |
| **Age** |  |  |  |  |  |  |
| Young | 547,387 | 62 | 0.37 | 0.54 (0.38-0.75) | 0.76 (0.53-1.09) | 0.132 |
| Old | 232,610 | 138 | 1.99 | 2.91 (2.21-3.85) | 1.59 (1.16-2.18) | 0.004 |
| Middle | 335,363 | 78 | 0.66 | 1 | 1 |  |
| **Economic status** |  |  |  |  |  |  |
| High | 350,615 | 87 | 0.78 | 1.00 (0.78-1.29) | 0.93 (0.72-1.20) | 0.571 |
| Low | 764,745 | 191 | 0.78 | 1 | 1 |  |
| **Region** |  |  |  |  |  |  |
| Metro | 592,778 | 145 | 0.78 | 1.00 (0.79-1.27) | 0.99 (0.78-1.26) | 0.943 |
| Rural | 522,582 | 133 | 0.77 | 1 | 1 |  |

**Supplementary Table. 3.**
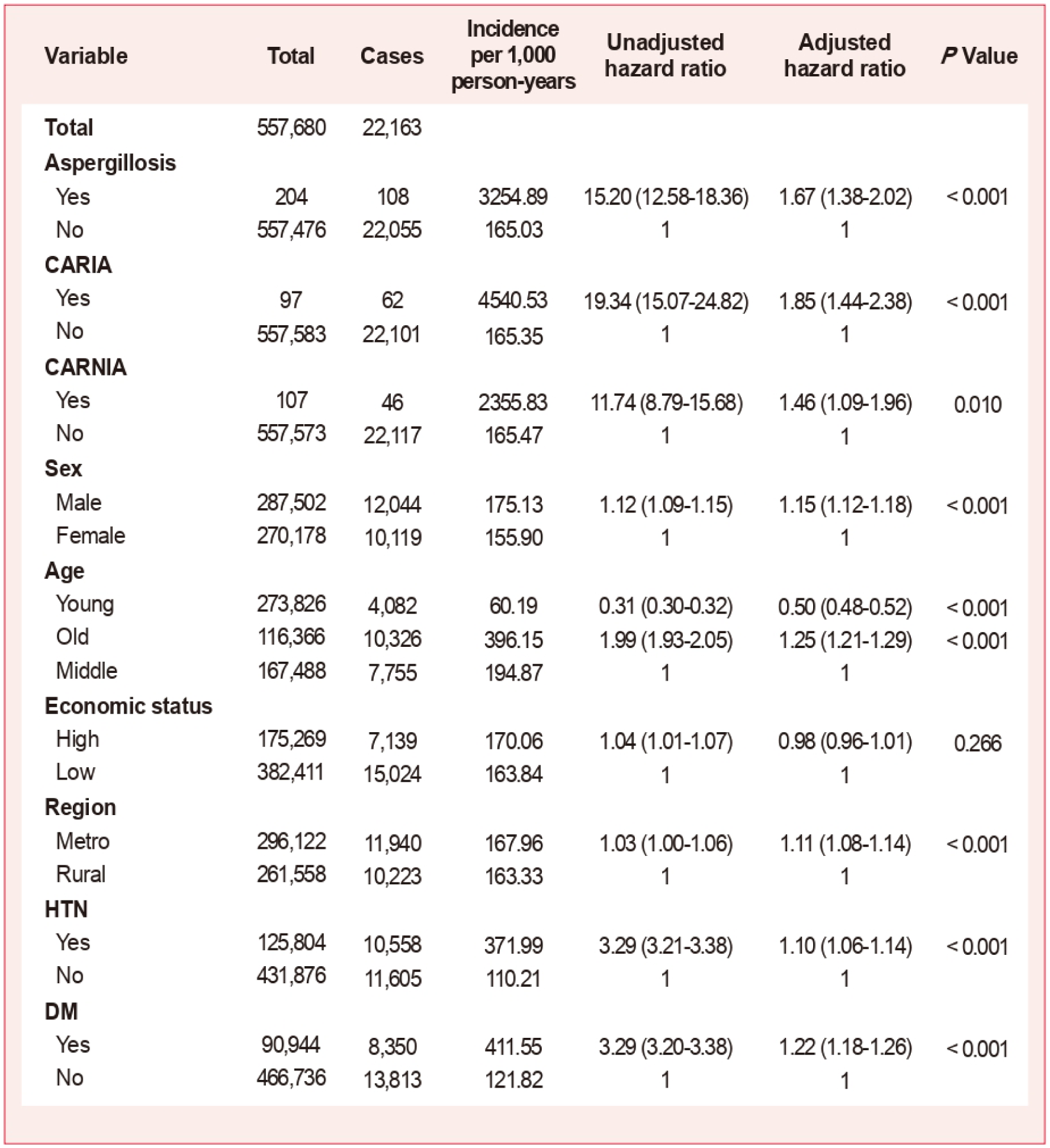

| Variable | Total | Cases | Incidence<br>per 1,000<br>person-years | Unadjusted<br>hazard ratio | Adjusted<br>hazard ratio | P Value |
| --- | --- | --- | --- | --- | --- | --- |
| **Total** | 557,680 | 22,163 |  |  |  |  |
| **Aspergillosis** |  |  |  |  |  |  |
| Yes | 204 | 108 | 3254.89 | 15.20 (12.58-18.36) | 1.67 (1.38-2.02) | < 0.001 |
| No | 557,476 | 22,055 | 165.03 | 1 | 1 |  |
| **CARIA** |  |  |  |  |  |  |
| Yes | 97 | 62 | 4540.53 | 19.34 (15.07-24.82) | 1.85 (1.44-2.38) | < 0.001 |
| No | 557,583 | 22,101 | 165.35 | 1 | 1 |  |
| **CARNIA** |  |  |  |  |  |  |
| Yes | 107 | 46 | 2355.83 | 11.74 (8.79-15.68) | 1.46 (1.09-1.96) | 0.010 |
| No | 557,573 | 22,117 | 165.47 | 1 | 1 |  |
| **Sex** |  |  |  |  |  |  |
| Male | 287,502 | 12,044 | 175.13 | 1.12 (1.09-1.15) | 1.15 (1.12-1.18) | < 0.001 |
| Female | 270,178 | 10,119 | 155.90 | 1 | 1 |  |
| **Age** |  |  |  |  |  |  |
| Young | 273,826 | 4,082 | 60.19 | 0.31 (0.30-0.32) | 0.50 (0.48-0.52) | < 0.001 |
| Old | 116,366 | 10,326 | 396.15 | 1.99 (1.93-2.05) | 1.25 (1.21-1.29) | < 0.001 |
| Middle | 167,488 | 7,755 | 194.87 | 1 | 1 |  |
| **Economic status** |  |  |  |  |  |  |
| High | 175,269 | 7,139 | 170.06 | 1.04 (1.01-1.07) | 0.98 (0.96-1.01) | 0.266 |
| Low | 382,411 | 15,024 | 163.84 | 1 | 1 |  |
| **Region** |  |  |  |  |  |  |
| Metro | 296,122 | 11,940 | 167.96 | 1.03 (1.00-1.06) | 1.11 (1.08-1.14) | < 0.001 |
| Rural | 261,558 | 10,223 | 163.33 | 1 | 1 |  |
| **HTN** |  |  |  |  |  |  |
| Yes | 125,804 | 10,558 | 371.99 | 3.29 (3.21-3.38) | 1.10 (1.06-1.14) | < 0.001 |
| No | 431,876 | 11,605 | 110.21 | 1 | 1 |  |
| **DM** |  |  |  |  |  |  |
| Yes | 90,944 | 8,350 | 411.55 | 3.29 (3.20-3.38) | 1.22 (1.18-1.26) | < 0.001 |
| No | 466,736 | 13,813 | 121.82 | 1 | 1 |  |

