## Supplementary figures and images for "Non-invasive aspergillosis following COVID-19 exacerbates the severity of SARS-CoV-2 infection"

### Supplementary figures and tables-01.png

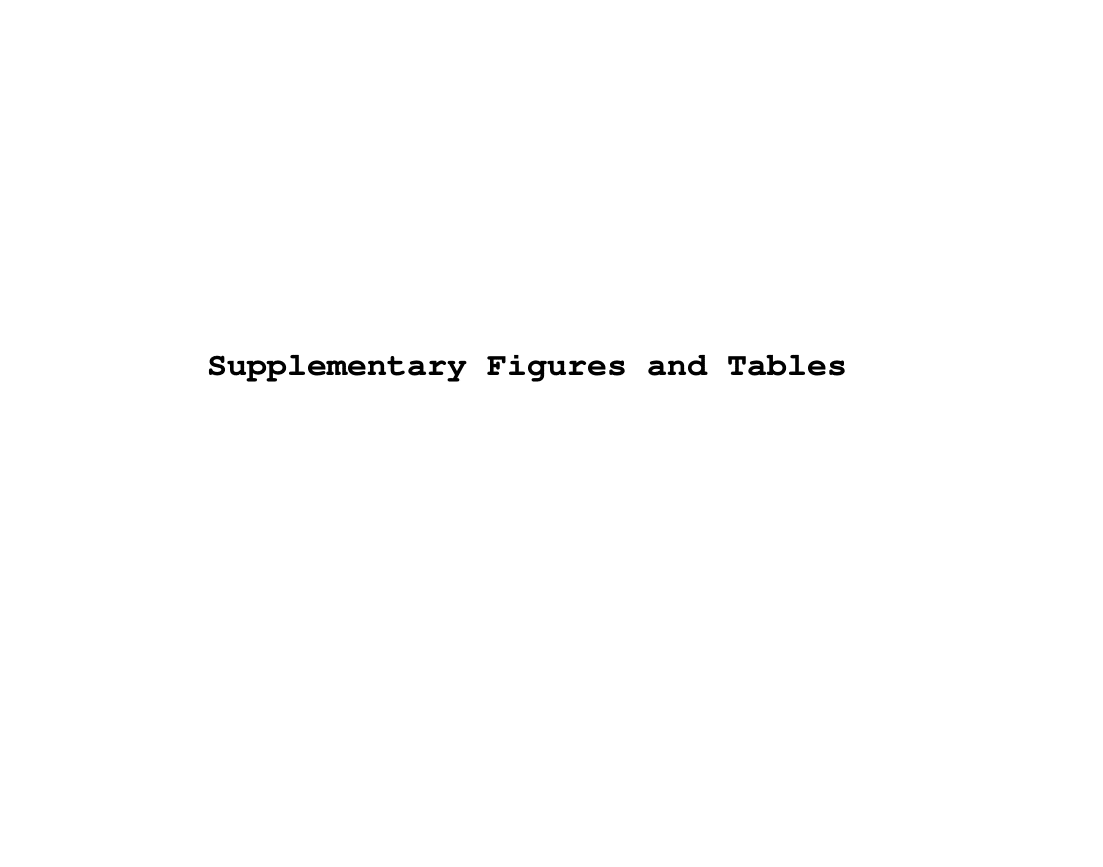

### Supplementary figures and tables-02.png

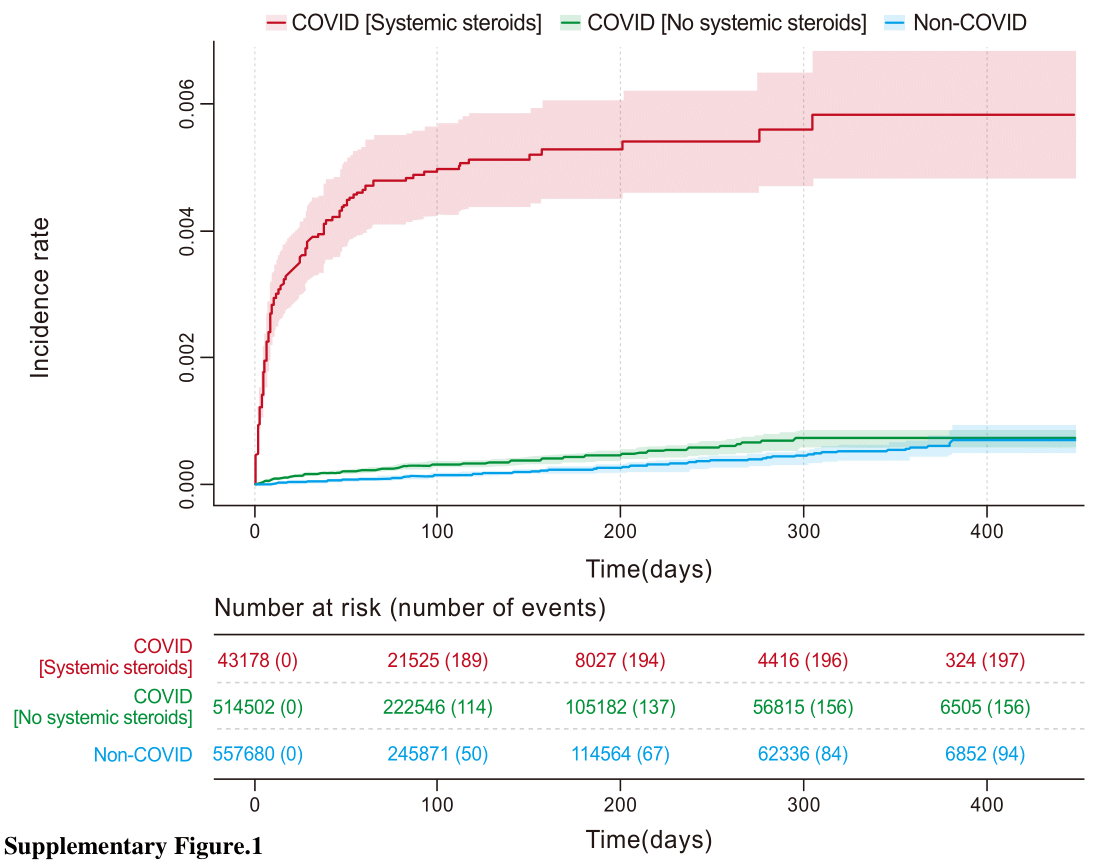

### Supplementary figures and tables-03.png

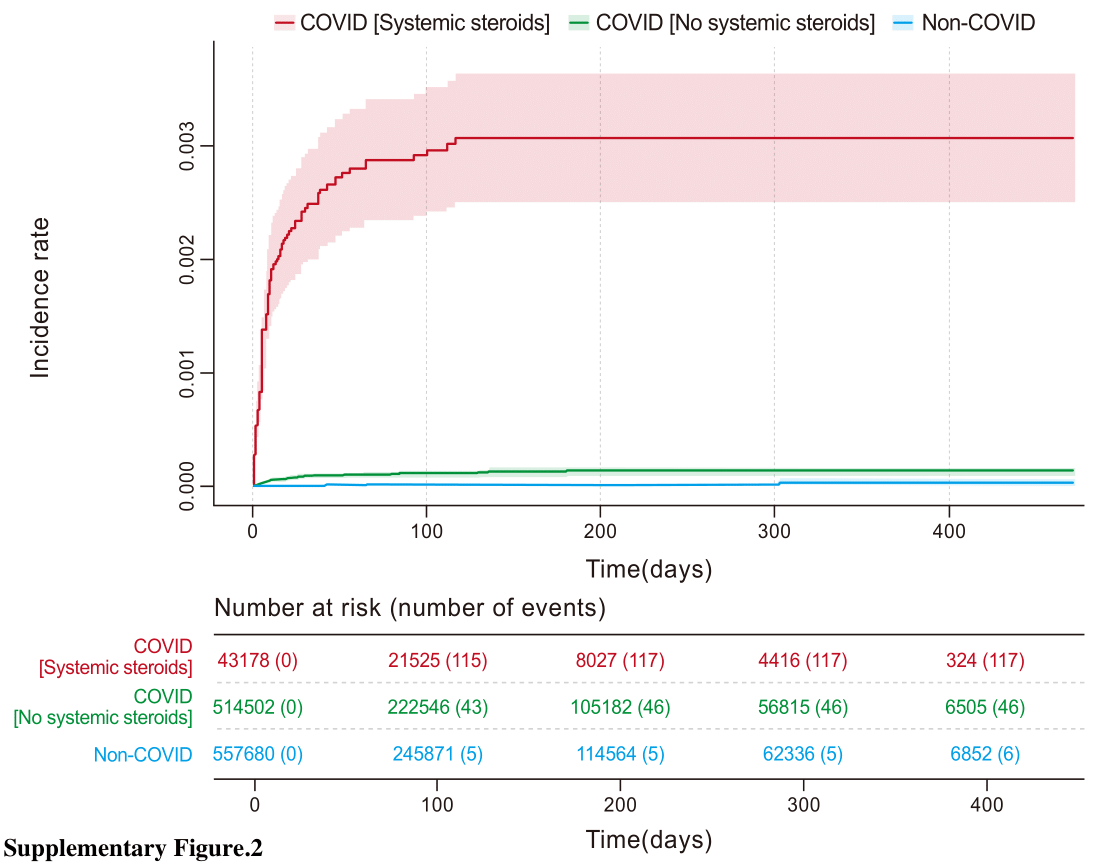

### Supplementary figures and tables-04.png

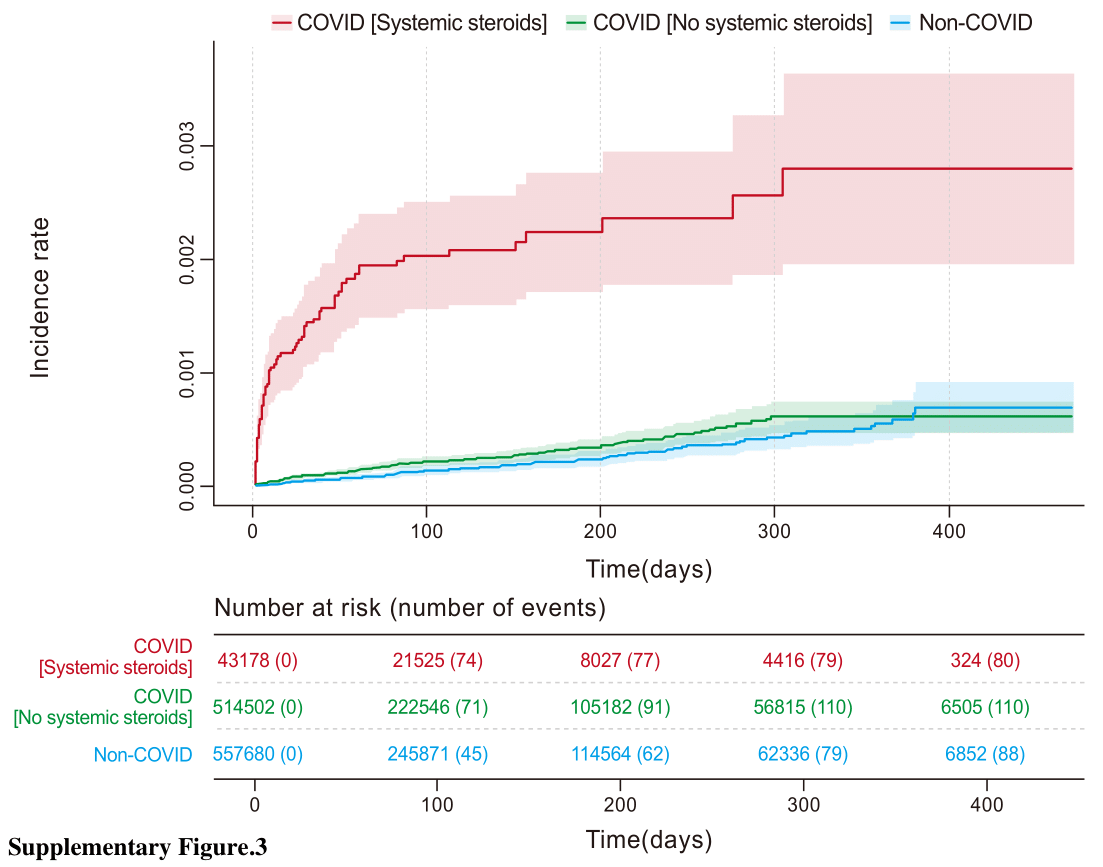

### Supplementary figures and tables-05.png

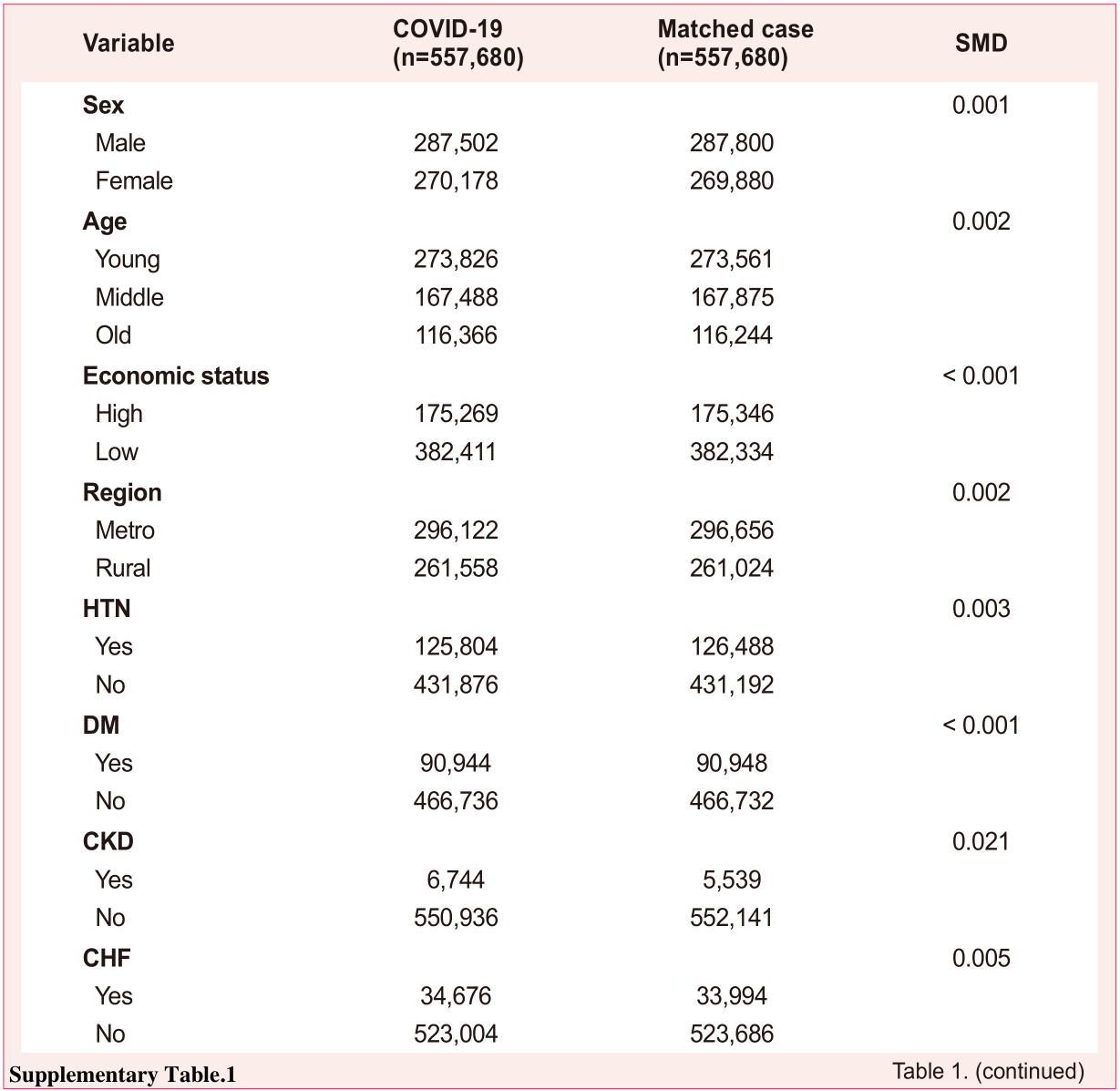

### Supplementary figures and tables-06.png

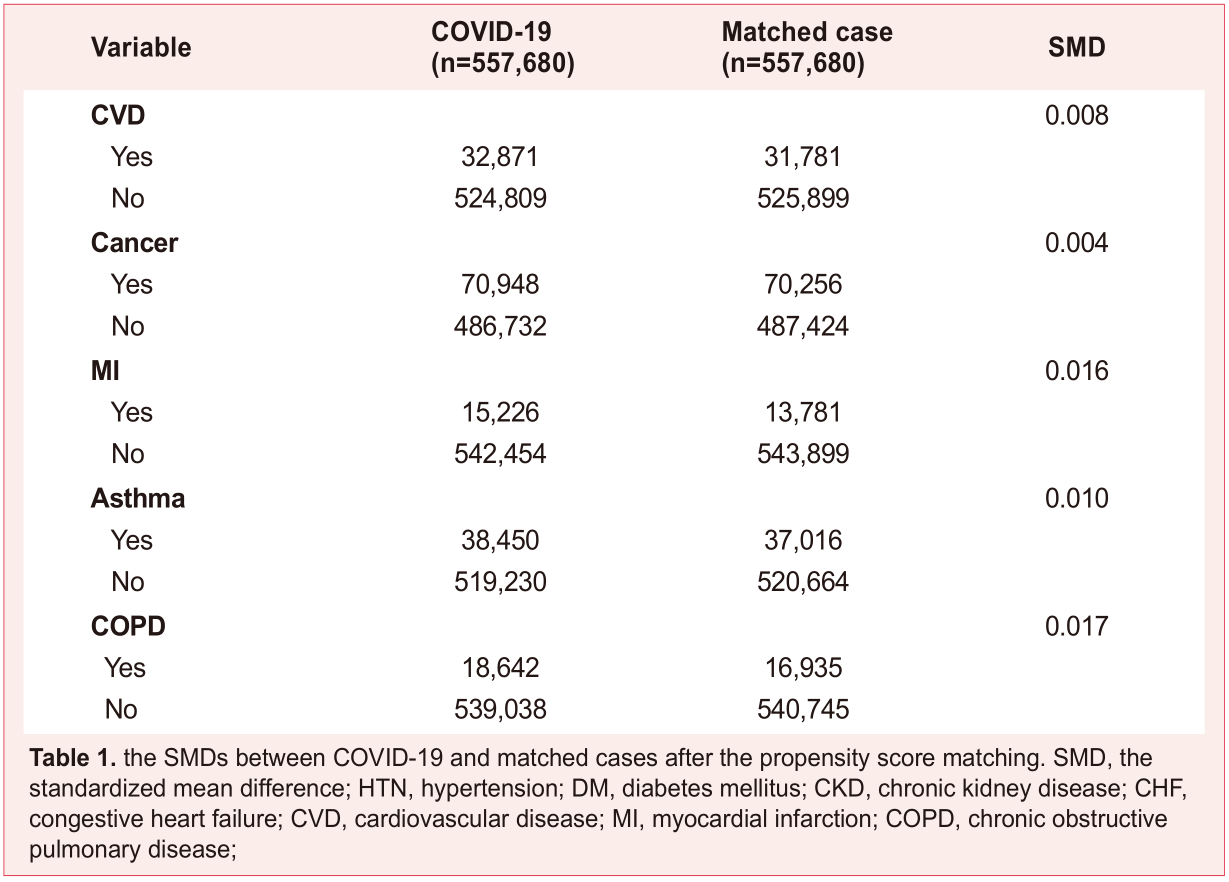

### Supplementary figures and tables-07.png

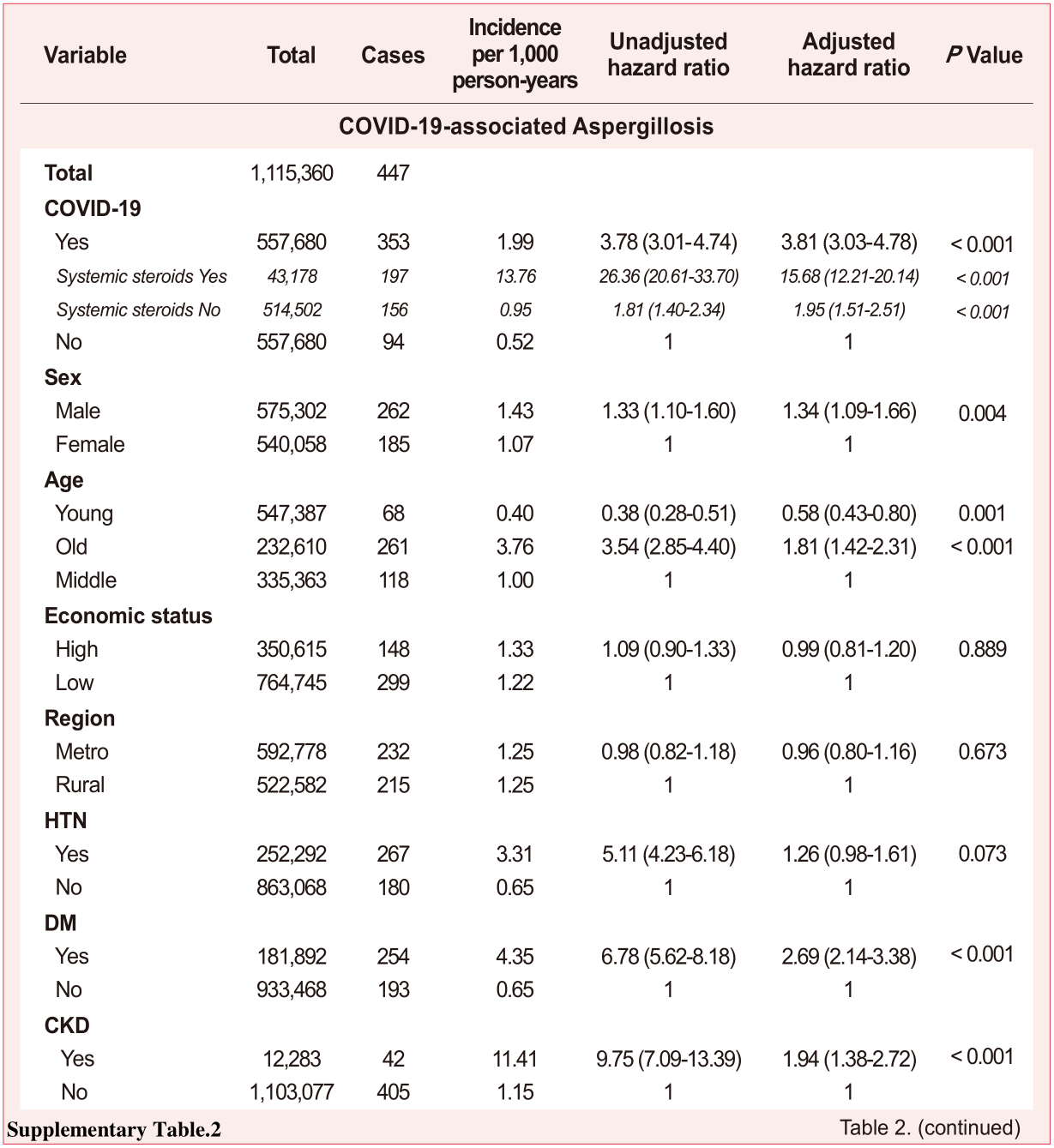

### Supplementary figures and tables-08.png

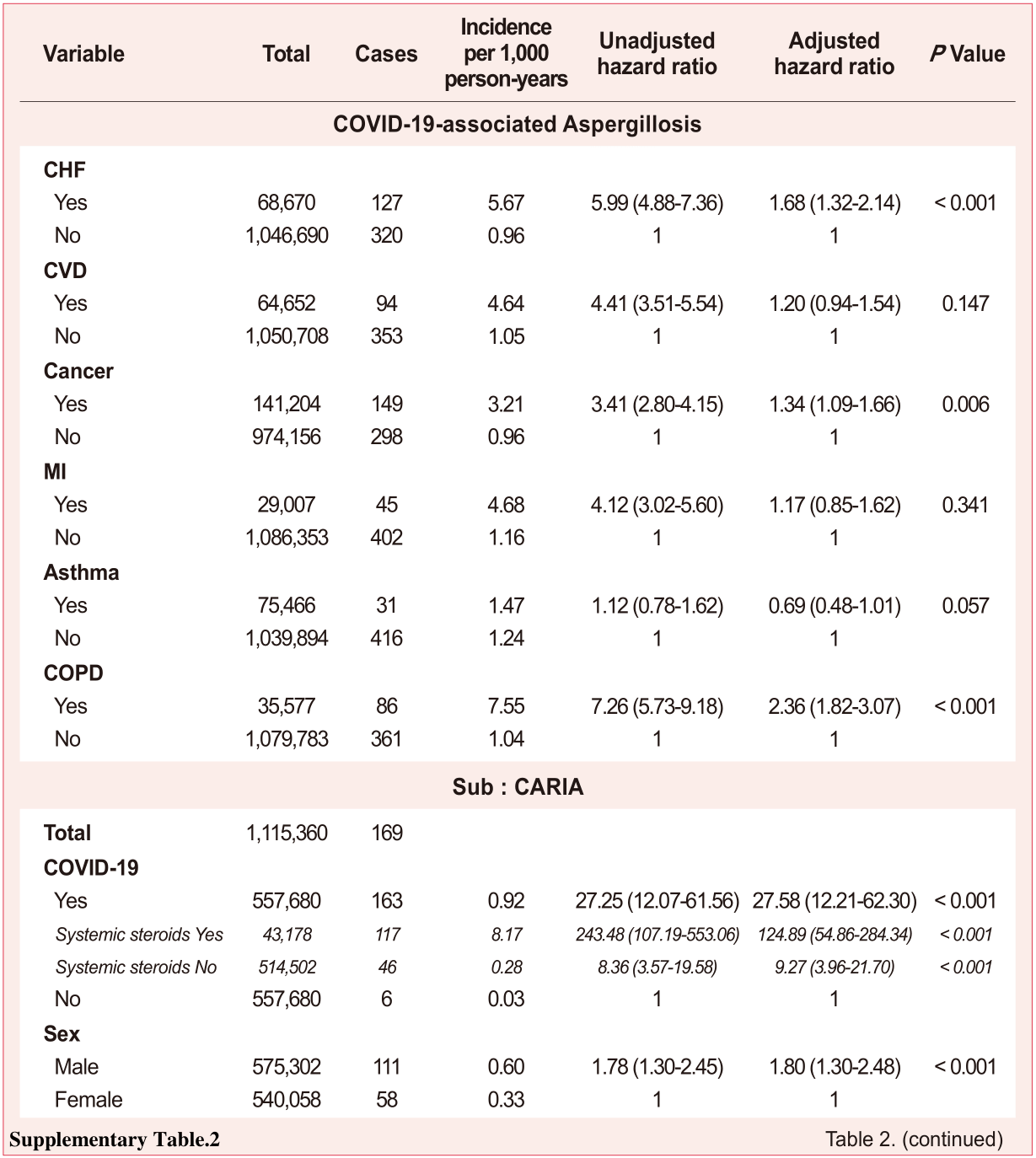

### Supplementary figures and tables-09.png

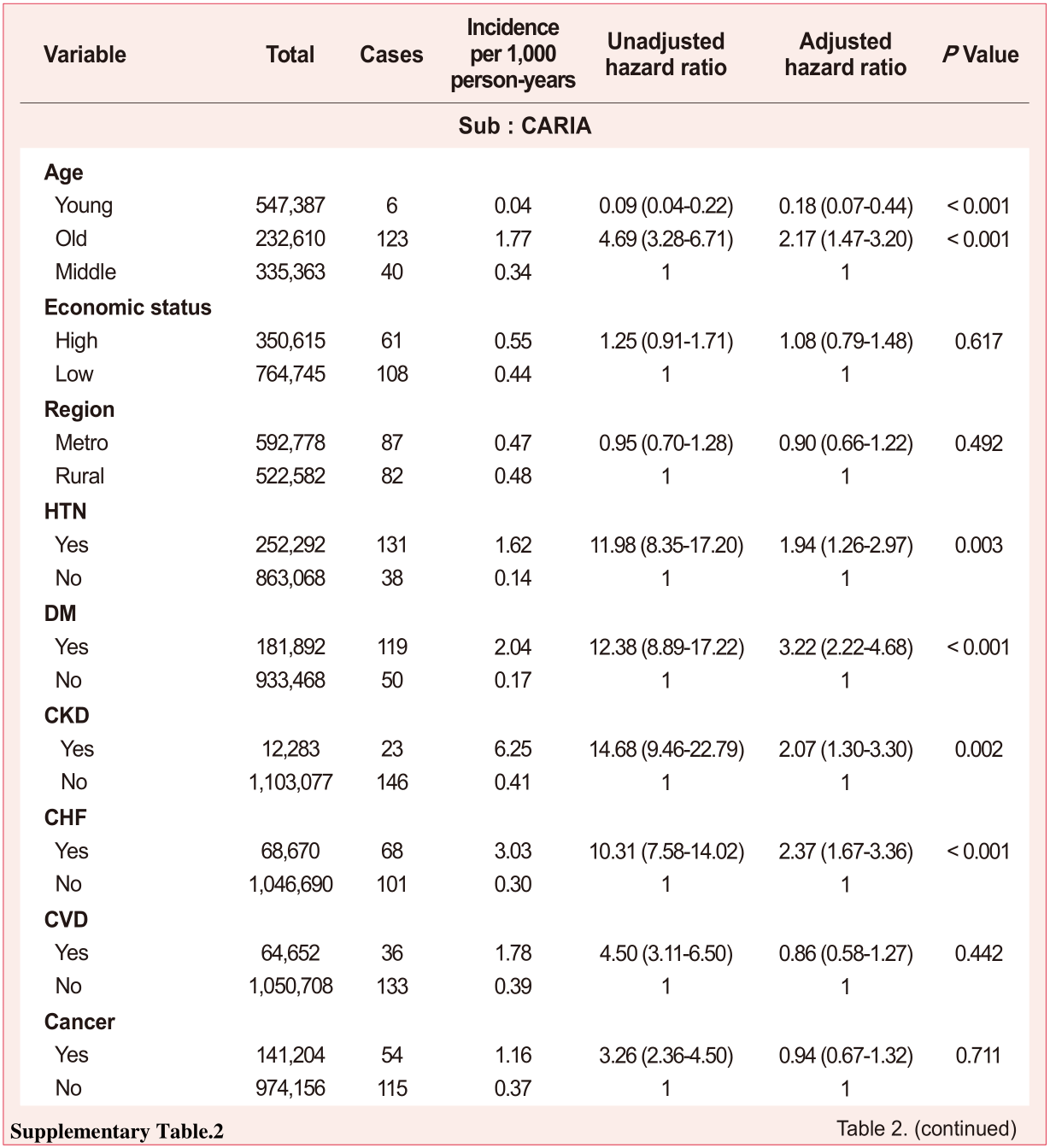

### Supplementary figures and tables-10.png

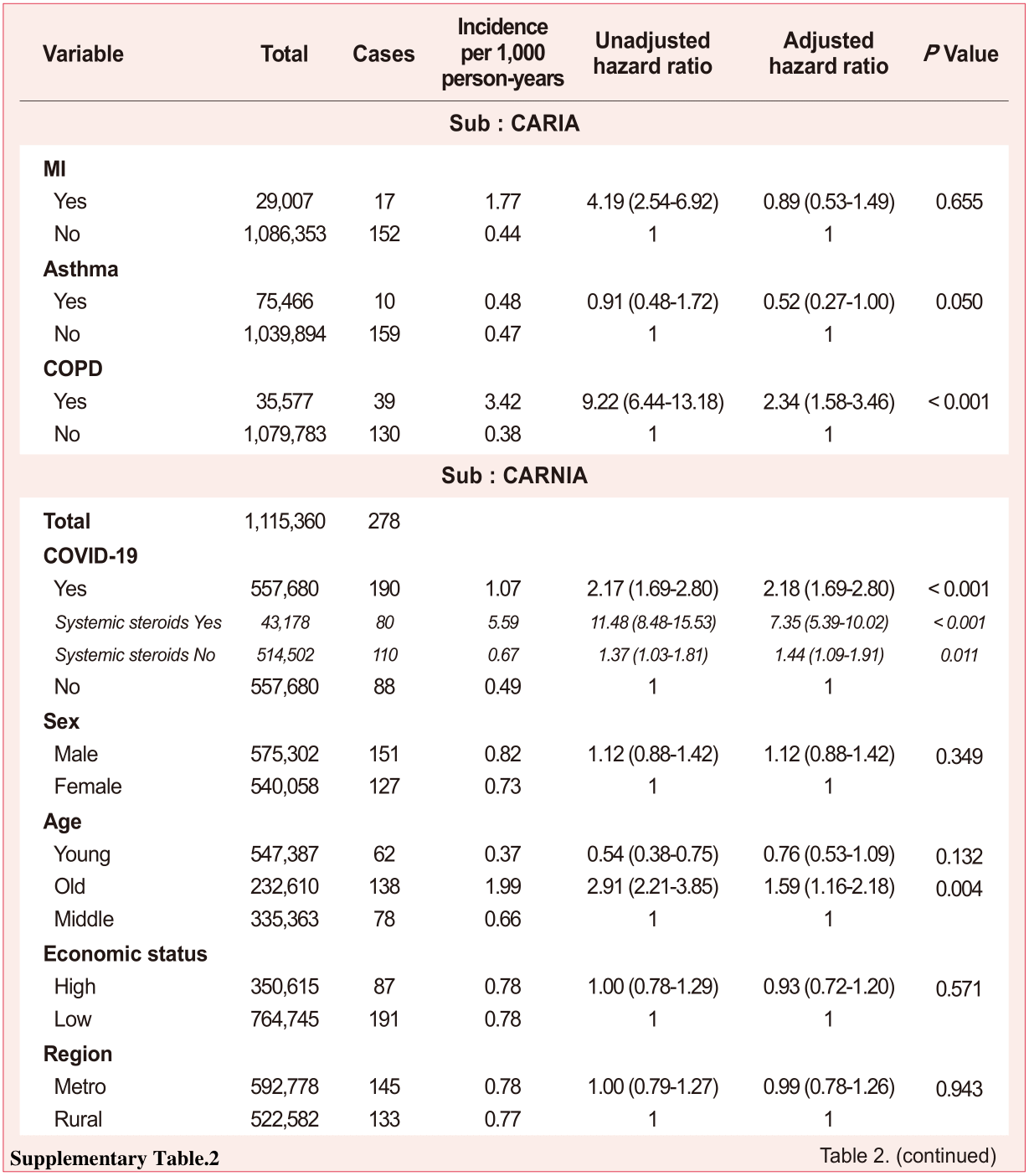

### Supplementary figures and tables-11.png

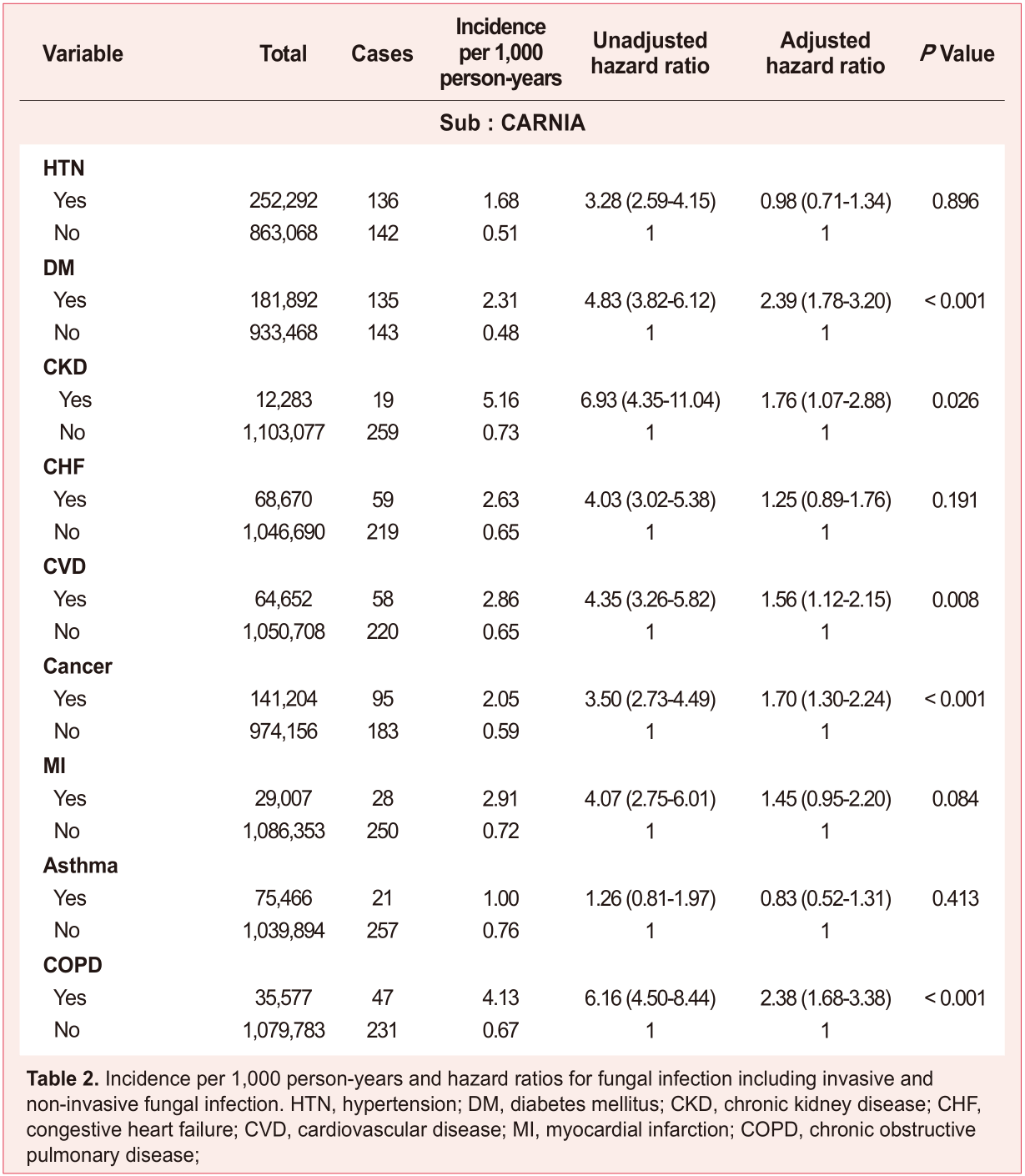

### Supplementary figures and tables-12.png

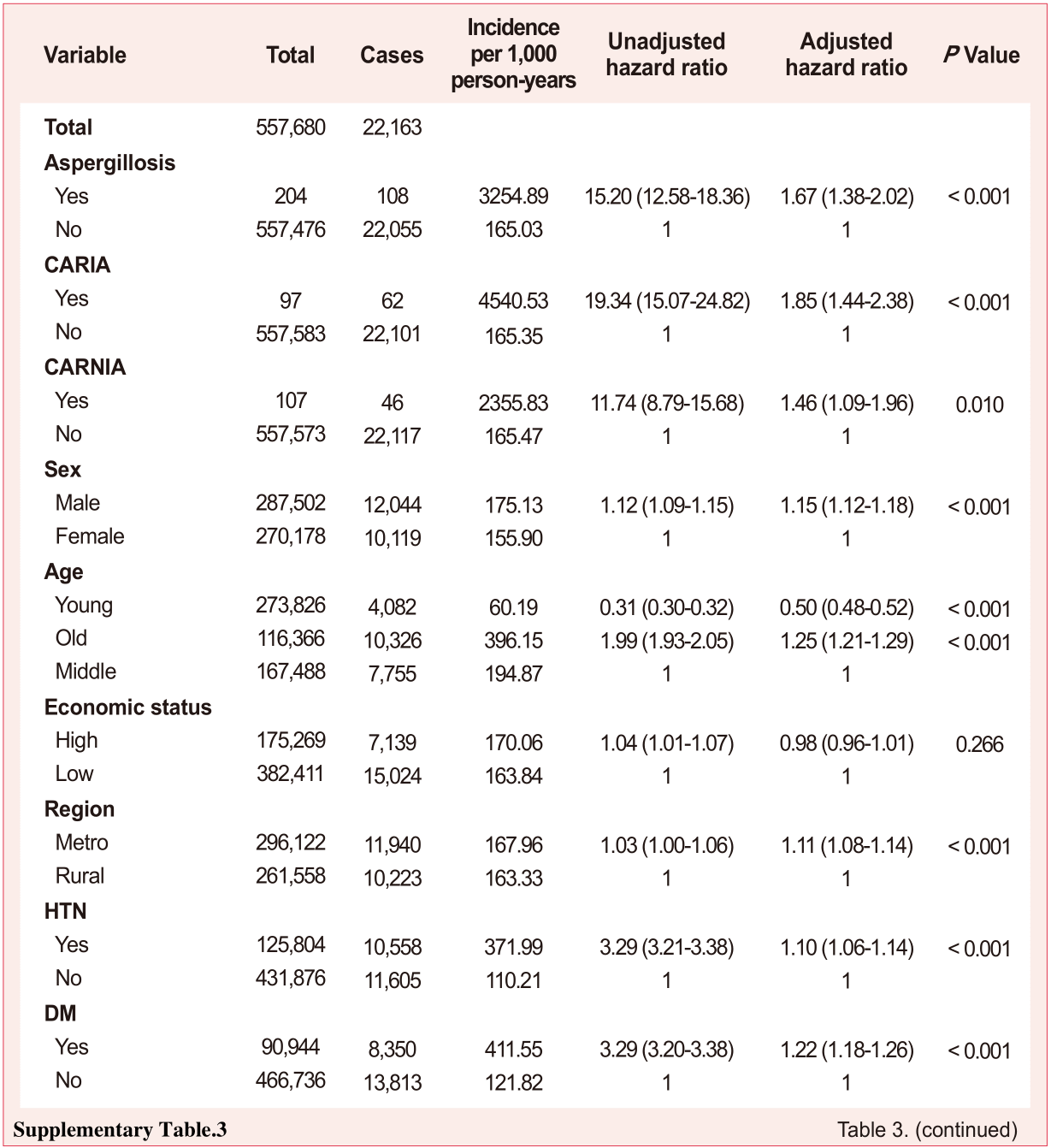

### Supplementary figures and tables-13.png

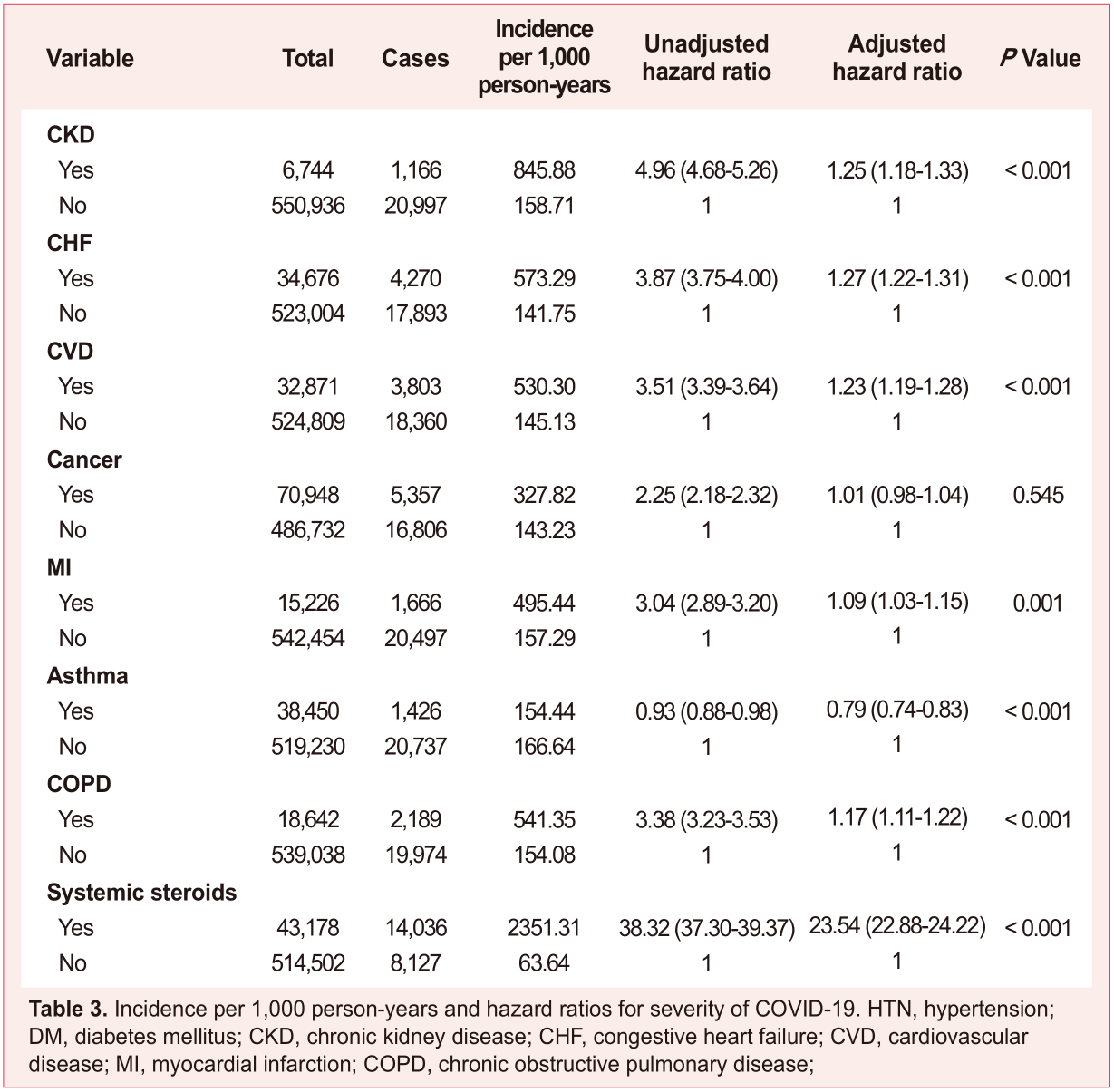
